# Persistent neuropsychiatric symptoms after COVID-19: a systematic review and meta-analysis

**DOI:** 10.1101/2021.04.30.21256413

**Authors:** James B. Badenoch, Emma R. Rengasamy, Cameron J. Watson, Katrin Jansen, Stuti Chakraborty, Ritika D. Sundaram, Danish Hafeez, Ella Burchill, Aman Saini, Lucretia Thomas, Benjamin Cross, Camille K. Hunt, Isabella Conti, Sylvia Ralovska, Zain Hussain, Matthew Butler, Thomas A. Pollak, Ivan Koychev, Benedict D. Michael, Heinz Holling, Timothy R. Nicholson, Jonathan P. Rogers, Alasdair G. Rooney, for the SARS-CoV-Neuro Collaboration

## Abstract

**Background:** The nature and extent of persistent neuropsychiatric symptoms after COVID-19 are not established. To help inform mental health service planning in the pandemic recovery phase, we systematically determined the prevalence of neuropsychiatric symptoms in survivors of COVID-19.

**Methods:** For this pre-registered systematic review and meta-analysis (PROSPERO ID CRD42021239750) we searched PubMed, EMBASE, CINAHL and PsycINFO to 20th February 2021, plus our own curated database. We included peer-reviewed studies reporting neuropsychiatric symptoms at post-acute or later time-points after COVID-19 infection, and in control groups where available. For each study a minimum of two authors extracted summary data. For each symptom we calculated a primary pooled prevalence using generalised linear mixed models. Heterogeneity was measured with *I*^2^. Subgroup analyses were conducted for COVID-19 hospitalisation, severity, and duration of follow-up.

**Findings:** From 2,844 unique titles we included 51 studies (*n*=18,917 patients). The mean duration of follow-up after COVID-19 was 77 days (range 14-182 days). Study quality was generally moderate. The most frequent neuropsychiatric symptom was sleep disturbance (pooled prevalence=27·4% [95%CI 21·4- 34·4%]), followed by fatigue (24·4% [17·5-32·9%]), objective cognitive impairment (20·2% [10·3-35·7%]), anxiety (19·1%[13·3-26·8%]), and post-traumatic stress (15·7% [9·9-24·1%]). Only two studies reported symptoms in control groups, both reporting higher frequencies in Covid-19 survivors versus controls. Between-study heterogeneity was high (I^2^=79·6%-98·6%). There was little or no evidence of differential symptom prevalence based on hospitalisation status, severity, or follow-up duration.

**Interpretation:** Neuropsychiatric symptoms are common and persistent after recovery from COVID-19. The literature on longer-term consequences is still maturing, but indicates a particularly high frequency of insomnia, fatigue, cognitive impairment, and anxiety disorders in the first six months after infection.

**Funding:** JPR is supported by the Wellcome Trust (102186/B/13/Z).

IK is funded through the NIHR (Oxford Health Biomedical Research Facility, Development and Skills Enhancement Award) and the Medical Research Council (Dementias Platform UK and Deep and Frequent Phenotyping study project grants).

HH is funded by the German Research Foundation (DFG, Grant: HO 1286/16-1). The funders played no role in the design, analysis or decision to publish.

**RESEARCH IN CONTEXT:** *Evidence before this study:* Neuropsychiatric symptoms like cognitive impairment, fatigue, insomnia, depression and anxiety can be highly disabling. Recently there has been increasing awareness of persistent neuropsychiatric symptoms after COVID-19 infection, but a systematic synthesis of these symptoms is not available. In this review we searched five databases up to 20th February 2021, to establish the pooled prevalence of individual neuropsychiatric symptoms up to six months after COVID-19.

*Added value of this study:* This study establishes which of a range of neuropsychiatric symptoms are the most common after COVID-19. We found high rates in general, with little convincing evidence that these symptoms lessen in frequency during the follow-up periods studied.

*Implications:* Persistent neuropsychiatric symptoms are common and appear to be limited neither to the post-acute phase, nor to recovery only from severe COVID-19. Our results imply that health services should plan for high rates of requirement for multidisciplinary services (including neurological, neuropsychiatric and psychological management) as populations recover from the COVID-19 pandemic.

## INTRODUCTION

Early in the COVID-19 pandemic, neuropsychiatric symptoms were identified as a prominent feature of coronavirus outbreaks. (1, 2) Analyses subsequently confirmed many neuropsychiatric manifestations of acute infection with SARS-CoV-2, with non-specific symptoms such as fatigue and headache the most commonly studied and reported in the early literature. (3) Studies assessing the prevalence of depression, anxiety, and post-traumatic stress in acute COVID-19 suggested specific psychiatric morbidity. The degree of persistence of neuropsychiatric symptoms in the post-acute and chronic phases after infection, however, remained far from clear.

Persistent symptoms after COVID-19 illness have been called “Long COVID”. (4–7) The point of onset of Long COVID is imprecisely defined and has been proposed to range from three to twelve weeks after infection. (8) Separately, UK guidelines conceptualise symptoms persisting between four and twelve weeks after infection as ‘ongoing symptomatic COVID-19’, with ‘post COVID-19 syndrome’ thereafter. (9) However it is defined, Long COVID is considered a multi-system disorder with most likely several distinct pathological mechanisms. (10–12) These uncertainties of definitions, terminology, and mechanism reflect the early stage of our knowledge about persisting symptoms after COVID-19, and in particular the lack of systematised descriptions of different components of the syndrome.

From a mental health perspective, emerging reports suggest a high frequency of neuropsychiatric symptoms after infection with COVID-19. These reports emphasise fatigue, cognitive dysfunction and sleep disorders, with increased rates of newly-diagnosed mood or anxiety disorders, and dementia. (13–15) Whether and how these neuropsychiatric sequelae are influenced by the severity of initial illness, or by the duration since COVID-19, is not known. Given the signals however, it seems likely that clinical services for COVID-19 survivors (8,16,17) will benefit from a synthesis of knowledge concerning the nature and extent of its neuropsychiatric complications. Previous analyses either did not focus specifically on these outcomes (18) or have been superseded by the rapid growth in research. (19)

We therefore aimed to estimate the prevalence of persistent neuropsychiatric symptoms in survivors of COVID-19. In secondary analyses we aimed to identify predictors of symptom prevalence. We hypothesised that persistent neuropsychiatric symptoms would be common among survivors of COVID- 19, particularly in those with a more severe form of the illness (i.e. those that have required hospitalisation or intensive care), and would lessen in frequency as time passed after infection.

## METHODS

We conducted a systematic review and meta-analysis based on a pre-registered protocol (PROSPERO ID CRD42021239750) reported according to PRISMA guidelines. (20) A detailed list of author contributions is provided (Table S1).

**Table 1.** Characteristics of included studies.

| Population characteristics (n = # of patients) |  |  |
| --- | --- | --- |
| Sample size (range, median) | n | % |
|  | 24-5879, 134 | - |
| COVID-19 confirmed by PCR | 15786/18917 | 83·4 |
| Age (mean, SD) | 50·9, 9·4 | - |
| Sex |  |  |
| Male | 6825 | 33·2 |
| Female | 6213 | 32·8 |
| Unstated | 5879 | 31·1 |
| Location of COVID-19 treatment |  |  |
| Hospital | 9970 | 52·7 |
| Community | 2957 | 15·7 |
| Mixed or Unstated | 5990 | 31·5 |
| COVID-19 Severity |  |  |
| ICU / WHO critical-sev | 1245 | 6·6 |
| Hospitalised unspecified | 8725 | 46·1 |
| Community only | 2957 | 15·6 |
| Mixed or Unstated | 5990 | 31·7 |
| Ethnicity |  |  |
| Stated | 2378 | 12·6 |
| Unstated | 16539 | 87·4 |
| Among stated |  |  |
| White | 1172 | 49·3 |
| Asian | 210 | 8·8 |
| Black | 182 | 7·7 |
| Mixed / multiple | 8 | 0·3 |
| Hispanic | 272 | 11·4 |
| Other (Arab or other) | 534 | 22·5 |
| Study Characteristics (n= # of studies) |  |  |
| Study design | n | % |
| Cohort | 43/51 | 84·3 |
| Cross-sectional | 8/51 | 15·7 |
| Has control group | 3/51 | 5·9 |
| Follow-up duration (range, mean) | 14-182, 77 | - |
| COVID19 time-point |  |  |
| <12 weeks | 30/51 | 58·8 |
| 12+ weeks | 19/51 | 37·3 |
| Unstated | 2/51 | 3·9 |
| Definition of follow-up duration |  |  |
| Post-discharge | 23/51 | 45·1 |
| Post-PCR & post-symptom onset | 16/51 | 31·4 |
| Other | 11/51 | 21·6 |
| Unspecified | 2/51 | 3·9 |

### Search strategy

We searched Ovid MEDLINE(R) and Epub Ahead of Print, In-Process and Other Non-Indexed Citations and Daily, EMBASE (via Ovid), APA PsycInfo (via OVID) and CINAHL (via EBSCO) from 1st January 2020 to 20th February 2021. We adapted a previously published, librarian-designed search strategy for post-acute, persisting, or Long COVID. (21) To maximise sensitivity, our search strategy (Supplemental Methods) did not specify neuropsychiatric terms. We further examined our weekly-curated database of COVID-19 neurology and neuropsychiatry research (22) for any papers that were missed by the search strategy, and screened the reference lists of relevant systematic reviews published at the time of our primary search. (18, 19)

### Eligibility criteria

We included any observational study reporting persistent neuropsychiatric symptoms in adults (aged 18+ years), with a history of PCR-confirmed or clinically suspected SARS-CoV-2 infection. We defined “persistence” of symptoms differently for hospitalised and community-based samples. In hospitalised samples, we considered persistent symptoms as those present after hospital discharge, because discharged individuals are generally beyond the acute illness phase. In community-based samples, which lacked a discharge date, we considered persistent symptoms as those still present at least four weeks after the onset of symptoms or a positive PCR test.

We adopted a definition of “neuropsychiatric” symptoms proposed by patient-led research in this area. (15) We studied: affective symptoms (specifically anxiety and panic attacks, depression, and mania); hallucinations; sleep disturbance; objectively reported cognitive impairment (i.e through standardised cognitive tests); subjective cognitive impairment (such as patient report of ‘brain fog’ or other lay terms); sensorimotor symptoms (such as paraesthesia, numbness, or weakness of specific body parts); dizziness and vertigo; headache; changes in speech or language; and changes in taste or smell. We added fatigue, which in our experience is commonly encountered in Long COVID clinics, and post-traumatic stress disorder or symptoms (PTSD/PTSS), which are frequently reported after COVID-19 (Box 1).

#### Box 1

##### Neuropsychiatric symptoms recorded in this review (adapted from Davis et al).

* Includes paraesthesia, numbness, or weakness of specific body parts

Anxiety

Depression

Mania

Sensorimotor*

Dizziness/vertigo

Sleep changes

Objectively measured cognitive dysfunction

Subjectively reported cognitive dysfunction

Headache

Reduced taste

Reduced smell

Speech or language difficulty

Hallucinations

Fatigue

Panic

Post-traumatic stress disorder/symptoms

We excluded studies which did not report original data; where patients were not infected (or presumed infected) with SARS CoV-2; had fewer than 10 COVID-19 patients; reported no post-discharge data (hospitalised samples) or no timepoints longer than 4 weeks post-diagnosis (community samples); did not report any of the neuropsychiatric symptoms listed above; were not in the English language; were preclinical (animal/laboratory-based); or had not been peer-reviewed. In addition and considering our main aim, studies were only eligible if their design permitted, in our opinion, a reasonably generalisable estimation of point prevalence to the wider population. On this basis for example we excluded studies in which participants were eligible solely because of predetermined characteristics (e.g., the presence of neurological symptoms), had been discharged to ongoing inpatient rehabilitation for persisting symptoms, or were primarily drawn from statistically enriched samples such as support groups designed for people with persisting symptoms. Senior authors (AR/JR) discussed and agreed decisions about eligibility taken on this basis (see also Results).

### Screening and data extraction

Screening of titles, abstracts, and full text was conducted by a minimum of two authors each blinded to the other’s ratings using Rayyan (www.rayyan.ai). Lead authors (AR/JR/JB) resolved assessment discrepancies. For each eligible study data were extracted to a customised spreadsheet by one reviewer, then checked for accuracy by a second reviewer.

We aimed to extract all usable data for primary and secondary analyses. For instance, if a study reported data on the total population plus a breakdown of the same data for individual subgroups, we extracted each group (total population, subgroup 1, subgroup 2, etc) separately to the database. In the primary analysis we only included data from the total population of each study. In order to qualify for a subgroup analysis, we took the conservative position that a study had to report extractable data on a completely homogeneous subgroup (e.g. we would not label studies reporting a combined 95% community and 5% hospitalised patients as ‘community’ samples; such a study would instead be excluded from the ‘hospital vs community’ secondary analysis). Where a study reported multiple time-points we included only the longest follow-up time-point. Where data were presented in a way which did not meet our purposes we contacted study investigators to request clarification. The quality of each study was graded using the Newcastle-Ottawa Scale by a minimum of two authors working independently. A full list of data fields and outcomes extracted is presented in Suppl Table 2.

**Table 2.** Pooled prevalence of individual neuropsychiatric symptoms (data for Fig 3).

| Symptom | N studies | N subjects | Pooled prevalence | 95% CI | I<sup>2</sup> |
| --- | --- | --- | --- | --- | --- |
| Fatigue | 32 | 7501 | 0·244 | [0·175, 0·329] | 98·0 % |
| Dysosmia | 18 | 4738 | 0·114 | [0·082, 0·156] | 93·1 % |
| Dysgeusia | 18 | 4675 | 0·074 | [0·047, 0·114] | 94·8 % |
| Depression | 16 | 10402 | 0·129 | [0·075, 0·215] | 98·6 % |
| Headache | 15 | 4023 | 0·066 | [0·036, 0·12] | 95·2 % |
| Anxiety | 14 | 3716 | 0·191 | [0·133, 0·268] | 95·8 % |
| Sleep problems | 12 | 4991 | 0·274 | [0·214, 0·344] | 95·6 % |
| Subj. cog. dysf. | 12 | 2336 | 0·153 | [0·089, 0·25] | 95·8 % |
| PTSD/PTSS | 9 | 2545 | 0·157 | [0·099, 0·241] | 95·3 % |
| Dizziness | 8 | 3665 | 0·029 | [0·016, 0·051] | 85·3 % |
| Obj. cog. dysf. | 6 | 727 | 0·202 | [0·103, 0·357] | 93·2 % |
| Sensorimotor | 5 | 607 | 0·055 | [0·024, 0·123] | 79·6 % |

### Outcomes

The primary outcome was the pooled prevalence of each neuropsychiatric symptom, using estimates of point prevalence where available. We recorded symptoms however defined or measured by study investigators, including on the basis of patient self-report, clinical interview, case-note review, or rating scale. Where rating scales were used to quantify symptoms, we noted the specific scale and threshold applied. Where symptoms were characterised as a dichotomous variable (present/absent), including for patients scoring above a rating scale threshold, we recorded the relative frequency (“n affected in study” divided by “n infected with SARS-CoV-2”). Where symptoms were reported as continuous or ordinal variables (e.g., using rating scales) *and* where such data were available for both COVID-19 patients and a control group, we intended to calculate and then pool the Standardised Mean Differences. Studies reporting continuous or ordinal variables in COVID-19 populations only were summarised narratively in a table.

### Primary analysis

We conducted the primary analysis on every neuropsychiatric symptom reported by three or more studies. We pooled results based on random-effects meta-analysis, using the metafor package in R version 4·0·2 (23) to calculate generalised linear mixed models for each prevalence outcome, (24, 25) before using the inverse variance method with the Freeman-Tukey double arcsine transformation as a comparative sensitivity analysis. (26) We assessed between-study heterogeneity using the I² statistic. For interpretation we reported forest plots with 95% confidence intervals.

Around one fifth of eligible studies reported multiple types of cognitive dysfunction. Post-hoc we separated cognitive dysfunction into ‘objective’ and ‘subjective’ dysfunction. We defined objective cognitive dysfunction as revealed by a cognitive assessment screening tool (e.g. MMSE, MoCA, or similar). All other forms of cognitive dysfunction (such as patient self-report of memory problems, ‘brain fog’, or similar) were classed as subjective. A small number of studies reported >1 symptom of subjective cognitive dysfunction (for example, self-reported “memory disorder” and “concentration disorder”, (27). In such cases we included the subjective cognitive symptom with the highest prevalence.

### Secondary analyses

A priori we intended to conduct secondary analyses examining for differences in neuropsychiatric symptom prevalence between a) COVID-19 patients and control groups, b) COVID-19 patients whose diagnosis was PCR-confirmed and those in whom it was not, c) hospitalised and community samples, and d) different time-points following a positive test for SARS-CoV-2 (specifying <12 weeks versus 12 or more weeks to align with a key timepoint in NICE guidance for post COVID-19 syndrome. (9) Following data extraction we discovered that analyses (a) and (b) could not be run owing to a dearth of studies with control groups or non-PCR-confirmed cases, and the wording of (d) was too restrictive (excluding for instance the many studies measuring duration of symptoms from the date of hospital discharge, rather than from the date of a positive test).

We added two post-hoc quantitative secondary analyses to reflect the balance of the literature. The first of these was a further evaluation of disease severity. We had found that some studies reported intensive care (ICU) vs non-ICU admission whereas others used the WHO severity scale. (28) We grouped these studies as follows: ICU admission OR reported as having WHO ‘critical’ or ‘severe’ COVID-19, versus hospitalised patients declared as not admitted to ICU. The second analysis was a broadening of the scope of ‘duration’ to include the 19 studies anchoring on discharge (<12 versus 12+ weeks). We did not combine studies anchoring on the date of PCR testing with those using the date of hospital discharge owing to the wide variability in duration of COVID-19 hospital admissions. However, post-hoc we did combine studies using the date of PCR testing with those using the date of onset of symptoms, which we reasoned were often likely to be close together in time. In a post-hoc qualitative analysis we inspected scatterplots of reported symptom prevalence against time (separately for: dichotomised <12/12+ weeks, mean duration, and median duration of follow-up for all symptoms).

We required a minimum of two studies in each sub-group being compared and a minimum of five eligible studies overall per analysis. Every secondary analysis conducted on each symptom is listed in Suppl Table 3.

**Table 3.** Pooled symptom prevalence by subgroups (data for Fig 4).

| Subgroup analysis | Symptom | Subgroup | N studies | N subjects | Pooled prevalence | 95% CI |
| --- | --- | --- | --- | --- | --- | --- |
| Hospitalisation status | Anxiety | hospitalised | 14 | 3555 | 0·187 | [0·131, 0·261] |
|  |  | non-hospitalised | 2 | 161 | 0·422 | [0·348, 0·500] |
|  | Depression | hospitalised | 14 | 3419 | 0·123 | [0·070, 0·207] |
|  |  | non-hospitalised | 2 | 560 | 0·088 | [0·052, 0·147] |
|  | Sleep problems | hospitalised | 10 | 4594 | 0·276 | [0·206, 0·358] |
|  |  | non-hospitalised | 2 | 161 | 0·366 | [0·296, 0·444] |
|  | Subj cog dysf | hospitalised | 8 | 1659 | 0·195 | [0·109, 0·325] |
|  |  | non-hospitalised | 2 | 199 | 0·100 | [0·049, 0·193] |
|  | Dizziness | hospitalised | 5 | 3220 | 0·038 | [0·021, 0·068] |
|  |  | non-hospitalised | 2 | 290 | 0·021 | [0·009, 0·045] |
|  | Headache | hospitalised | 10 | 3049 | 0·051 | [0·024, 0·106] |
|  |  | non-hospitalised | 2 | 283 | 0·106 | [0·075, 0·148] |
|  | Fatigue | hospitalised | 23 | 5112 | 0·268 | [0·183, 0·375] |
|  |  | non-hospitalised | 3 | 386 | 0·214 | [0·156, 0·286] |
|  | PTSD/PTSS | hospitalised | 9 | 1926 | 0·154 | [0·098, 0·233] |
|  |  | non-hospitalised | 3 | 619 | 0·153 | [0·060, 0·337] |
| Severity | Anxiety | ITU / critical / severe | 3 | 220 | 0·414 | [0·241, 0·611] |
|  |  | non-ITU | 3 | 443 | 0·214 | [0·088, 0·437] |
|  | Sleep problems | ITU / critical / severe | 4 | 264 | 0·292 | [0·24, 0·349] |
|  |  | non-ITU | 3 | 1814 | 0·260 | [0·241,<br>0·281] |
|  | Subj cog<br>dysf· | ITU / critical /<br>severe | 3 | 83 | 0·277 | [0·192,<br>0·383] |
|  |  | non-ITU | 3 | 271 | 0·316 | [0·211,<br>0·444] |
|  | Headache | ITU / critical /<br>severe | 2 | 184 | 0·016 | [0·005,<br>0·049] |
|  |  | non-ITU | 2 | 1680 | 0·018 | [0·013,<br>0·025] |
|  | Fatigue | ITU / critical /<br>severe | 6 | 383 | 0·335 | [0·102,<br>0·692] |
|  |  | non-ITU | 5 | 625 | 0·246 | [0·053,<br>0·655] |
|  | Dysgeusia | ITU / critical /<br>severe | 3 | 173 | 0·075 | [0·044,<br>0·125] |
|  |  | non-ITU | 3 | 1814 | 0·073 | [0·062,<br>0·086] |
|  | Dysosmia | ITU / critical /<br>severe | 4 | 264 | 0·098 | [0·068,<br>0·141] |
|  |  | non-ITU | 3 | 1814 | 0·104 | [0·091,<br>0·119] |
| Time since discharge | Anxiety | <12 weeks | 10 | 2537 | 0·170 | [0·116,<br>0·242] |
|  |  | 12+weeks | 2 | 672 | 0·200 | [0·040,<br>0·601] |
|  | Depression | <12 weeks | 9 | 2304 | 0·118 | [0·051,<br>0·252] |
|  |  | 12+weeks | 3 | 710 | 0·147 | [0·045,<br>0·387] |
|  | Sleep<br>problems | <12 weeks | 3 | 674 | 0·362 | [0·241,<br>0·504] |
|  |  | 12+weeks | 4 | 2496 | 0·259 | [0·199,<br>0·330] |
|  | Headache | <12 weeks | 4 | 624 | 0·066 | [0·013,<br>0·280] |
|  |  | 12+weeks | 4 | 2086 | 0·029 | [0·004,<br>0·178] |
|  | Fatigue | <12 weeks | 11 | 2817 | 0·254 | [0·101,<br>0·506] |
|  |  | 12+weeks | 7 | 1074 | 0·268 | [0·161,<br>0·412] |
|  | Dysgeusia | <12 weeks | 3 | 334 | 0·048 | [0·015,<br>0·147] |
|  |  | 12+weeks | 5 | 2220 | 0·066 | [0·057,<br>0·077] |
|  | Dysosmia | <12 weeks | 3 | 334 | 0·061 | [0·025,<br>0·141] |
|  |  | 12+weeks | 5 | 2256 | 0·080 | [0·056,<br>0·112] |
|  | PTSD/PTSS | <12 weeks | 5 | 1202 | 0·179 | [0·116,<br>0·266] |
|  |  | 12+weeks | 2 | 358 | 0·180 | [0·036,<br>0·563] |
| Time since symptom<br>onset | Headache | <12 weeks | 4 | 571 | 0·115 | [0·051,<br>0·238] |
|  |  | 12+weeks | 3 | 742 | 0·066 | [0·050,<br>0·086] |
|  | Fatigue | <12 weeks | 5 | 1015 | 0·235 | [0·125,<br>0·400] |
|  |  | 12+weeks | 5 | 634 | 0·197 | [0·139,<br>0·272] |
|  | Dysgeusia | <12 weeks | 2 | 254 | 0·049 | [0·014,<br>0·163] |
|  |  | 12+weeks | 5 | 908 | 0·105 | [0·058,<br>0·182] |
|  | Dysosmia | <12 weeks | 2 | 254 | 0·177 | [0·135,<br>0·230] |
|  |  | 12+weeks | 5 | 935 | 0·158 | [0·118,<br>0·208] |

## RESULTS

### Study selection

The search yielded 4385 studies. After de-duplication we screened the titles and abstracts of 2861 studies, the full text of 428 studies, and included 51 eligible studies (Figure 1, Table S4 (27,29–78). Brief reasons for excluding studies are listed in Table S5. We contacted the authors of seven studies which did not report extricable prevalence data; three replied with usable data.

**Figure 1.**
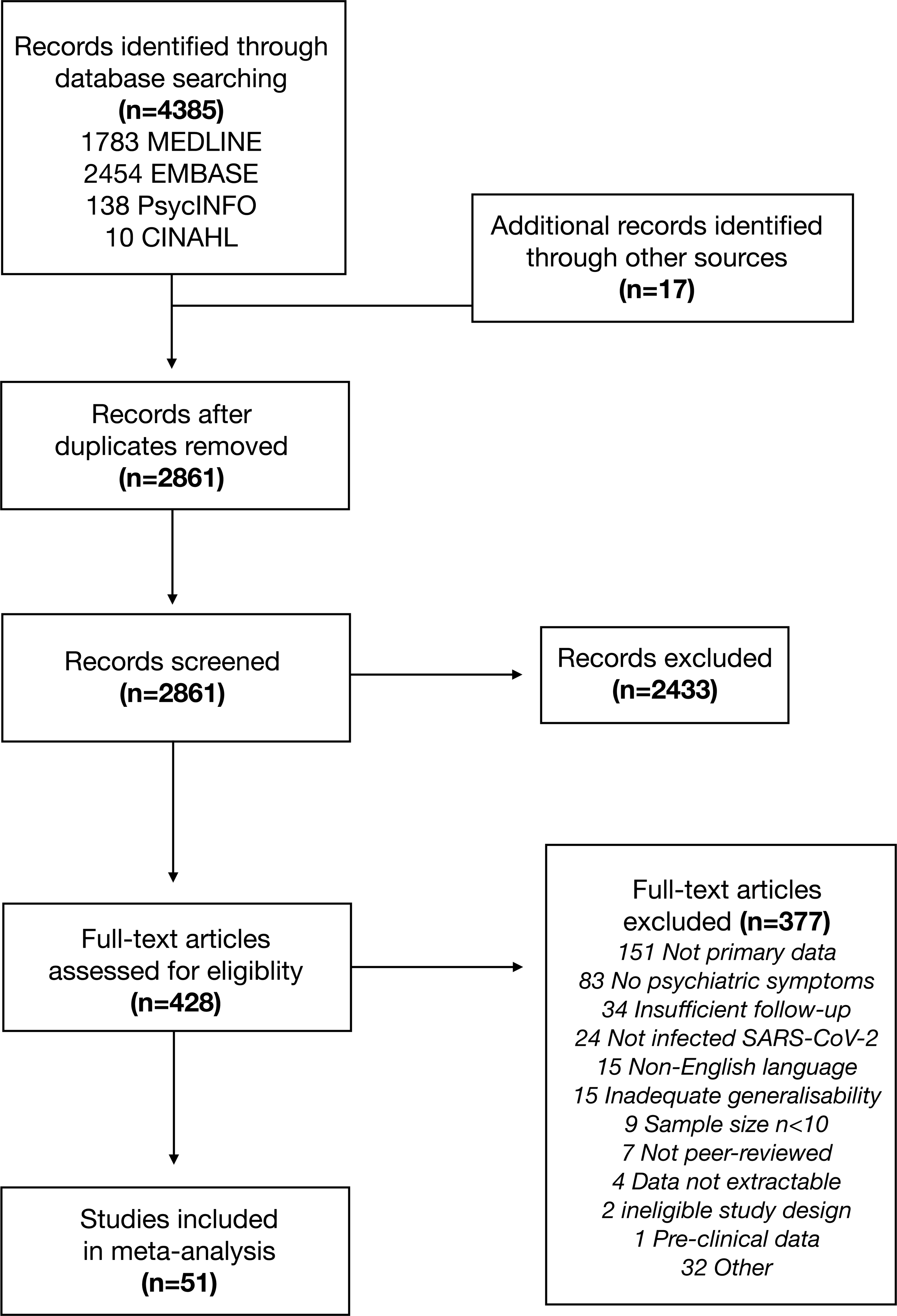
PRISMA Flowchart.

### Population and study characteristics

The 51 included studies reported data on a total population *n*=18917 individuals (sample size range 24- 5879; median *n*=134), *n*=15786 (83·4%) of whom had COVID-19 confirmed by PCR. The mean age reported was 50·9 years (SD=9·4). The largest study (*n*=5879, 31·1% of entire sample) did not specify sex; of the remainder *n*=6825 (33·2%) of patients were specified male. Most patients (*n*=9970, 52·8%) were post-hospital discharge, *n*=2957 (15·7%) had been treated solely in the community, and in *n*=5962 (31·5%) the location of care was unstated. A metric of COVID-19 severity was reported by half (26/51) of studies but the number of severe cases declared was often small: only *n*=1245/18917 (6·6%) of patients in the full sample were specified as having had ICU admission or WHO critical/severe COVID-19.

Ethnicity was reported for only *n*=2378/18917 (12·6%) patients, in whom *n*=1172 (49·3% of specified) were White. Most studies originated in China (13 studies) followed by Italy (six studies), the USA (5 studies), and the UK (5 studies) (Figure 2, Table 1).

**Figure 2.**
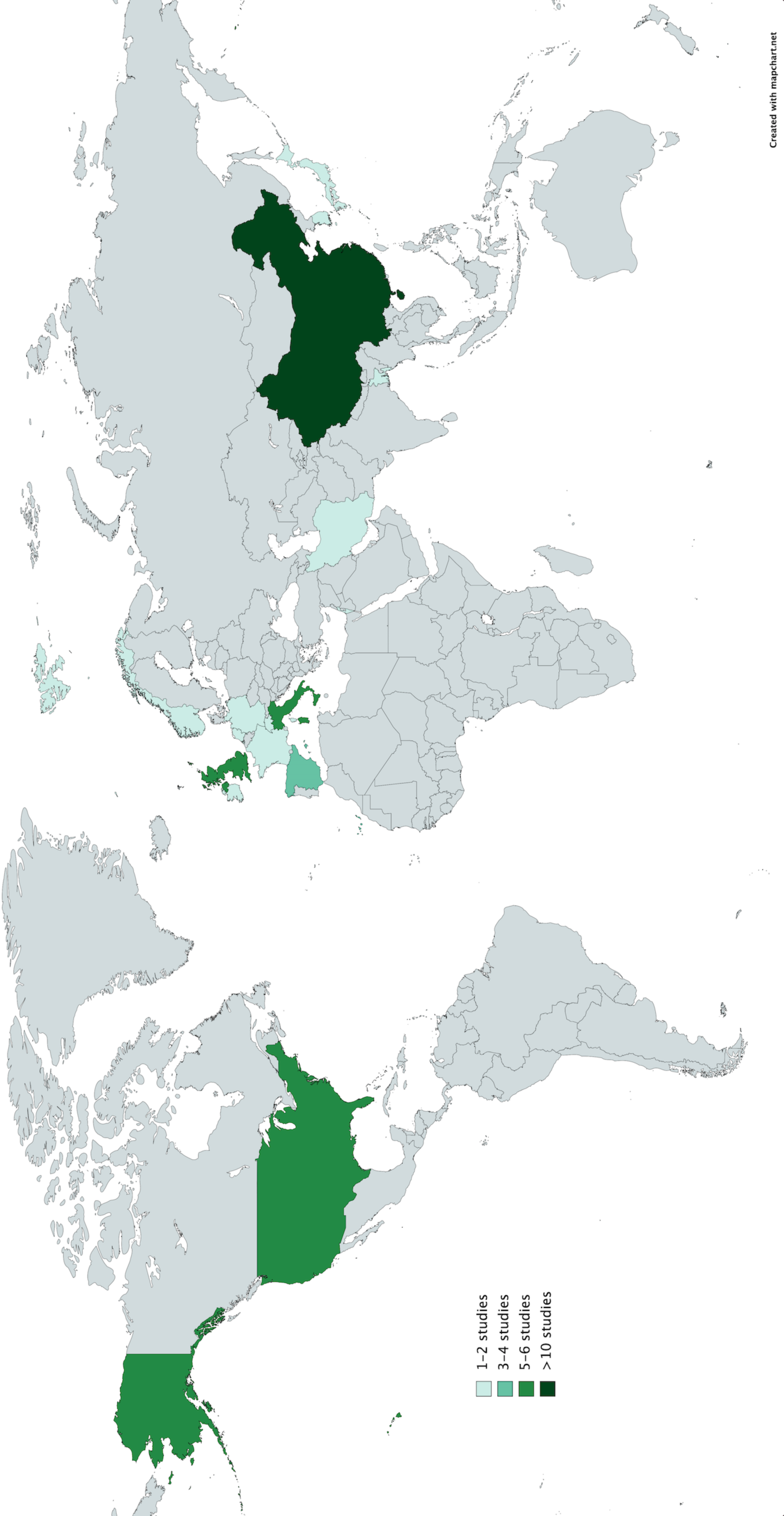
Map of studies. Most studies originated in China, the USA, the UK, and Italy. Map created with mapchart.net.

Most studies (43/51) had a cohort design. Only 2/51 reported symptoms in a control group. The mean duration of followup was 77 days (range 14-182 days). Most studies (30/51) examined patients at <12 weeks with 19/51 examining a time-point at 12+ weeks. There was little consensus on the anchor point for calculating follow-up duration: 23 studies reported duration since hospital discharge, 16 studies reported duration post-symptom onset or post-PCR testing, 11 used another definition (e.g. date of virological clearance), and two did not specify the anchor point. Study quality assessed using the Newcastle-Ottawa Scale was low in seven studies, medium in 39, and high in five (Figure S1, Table S6).

### Prevalence of neuropsychiatric symptoms

The most frequent neuropsychiatric symptom was sleep disturbance (pooled prevalence=27·4% [95%CI 21·4-34·4%]), followed by fatigue (24·4% [17·5-32·9%]), objectively measured cognitive impairment (20·2% [10·3-35·7%]), anxiety (19·1% [13·3-26·8%]), and post-traumatic stress (15·7% [9·9-24·1%]) (Figure 3, Table 2). More classically ‘neurological’ symptoms such as dysgeusia, headache, sensorimotor disturbance, and dizziness/vertigo were less frequent but present in non-negligible amounts (pooled prevalence <10% for each). Speech and language symptoms, panic attacks, mania, and hallucinations could not be meta-analysed due to an absence of studies. Heterogeneity was high (*I*^2^=79·6%-98·6%, Table 2). The results of the sensitivity analysis were in general similar to the results of the main analysis in terms of the point estimate of prevalence, confidence interval boundaries and heterogeneity (Table S7).

**Figure 3.**
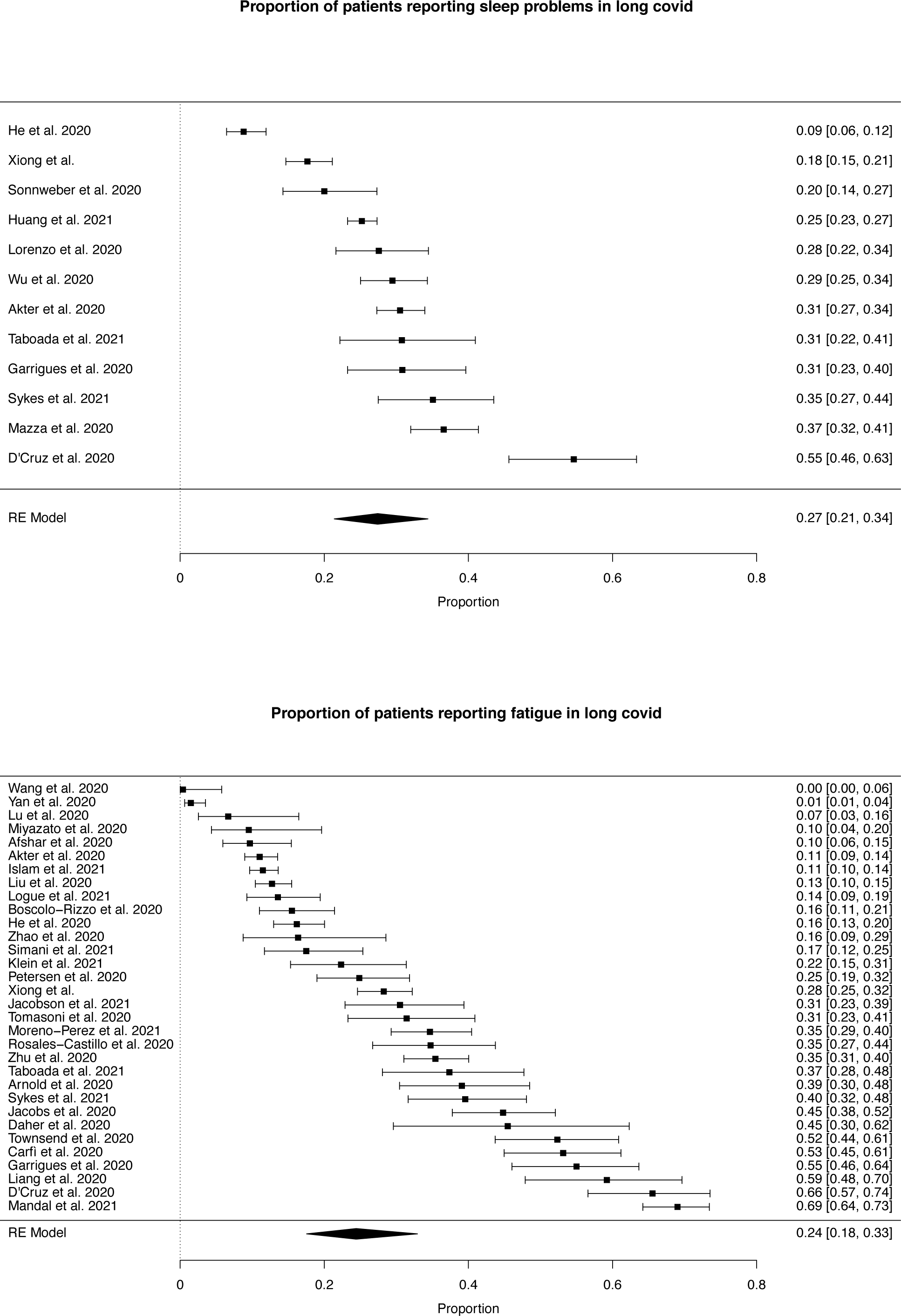
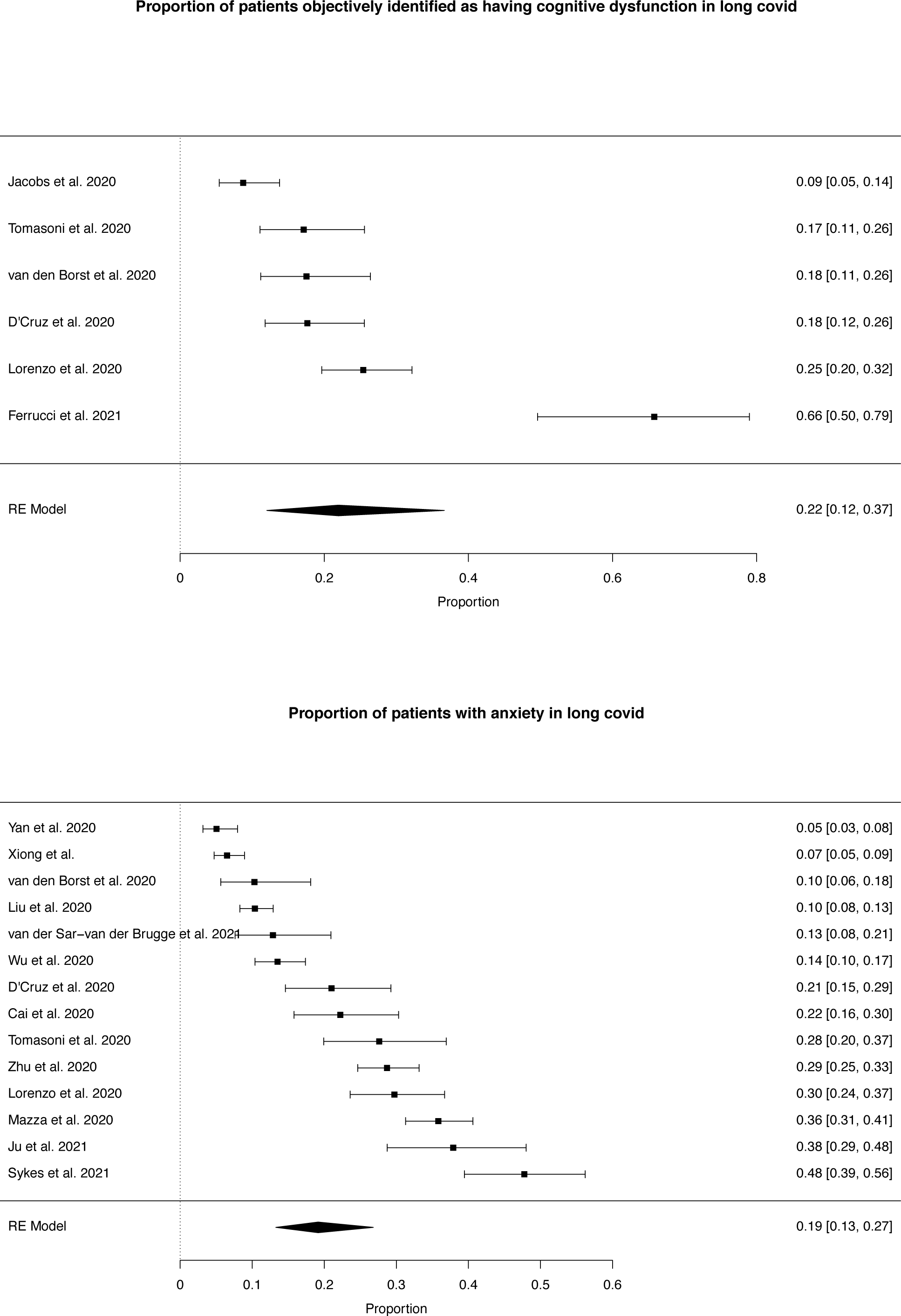
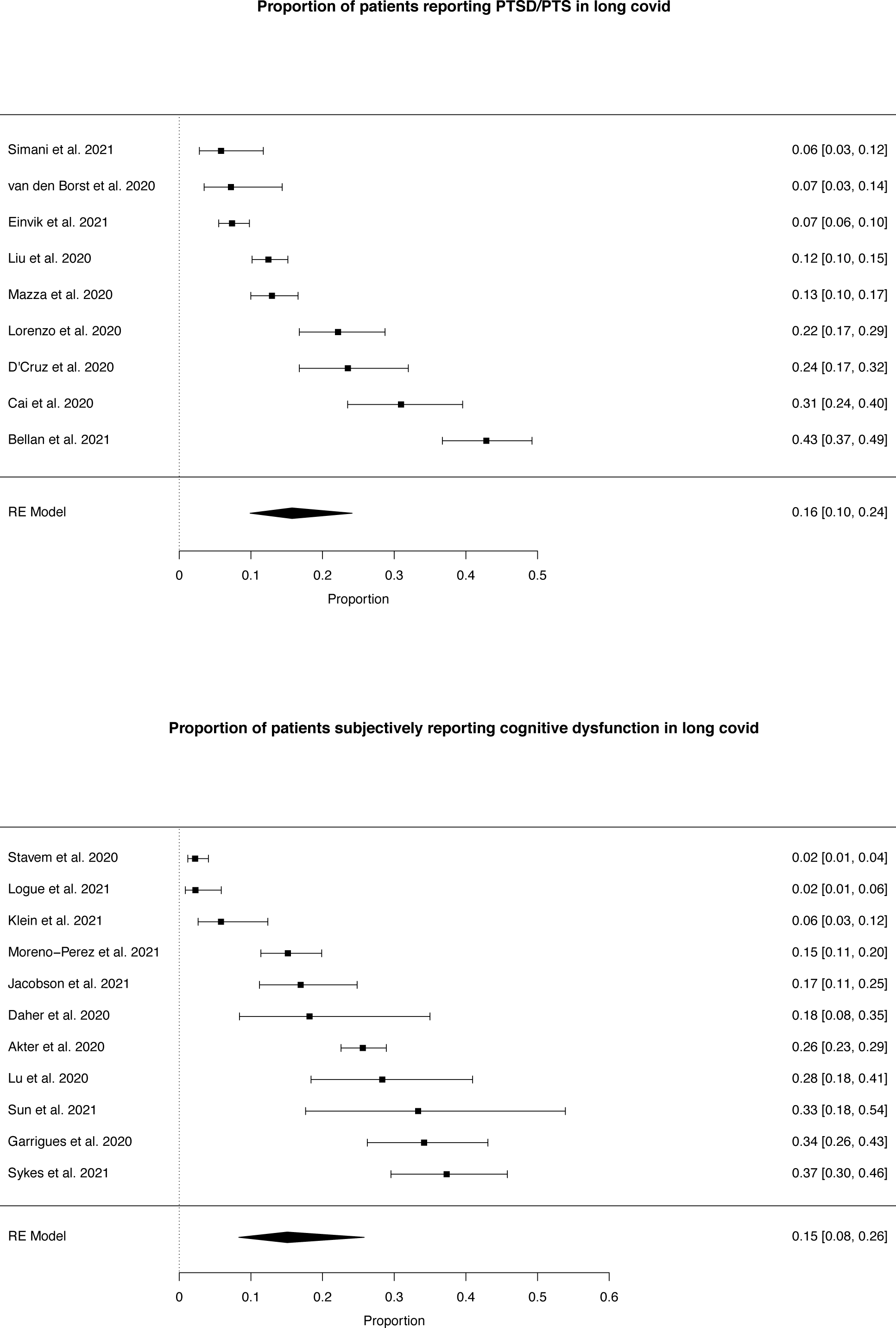
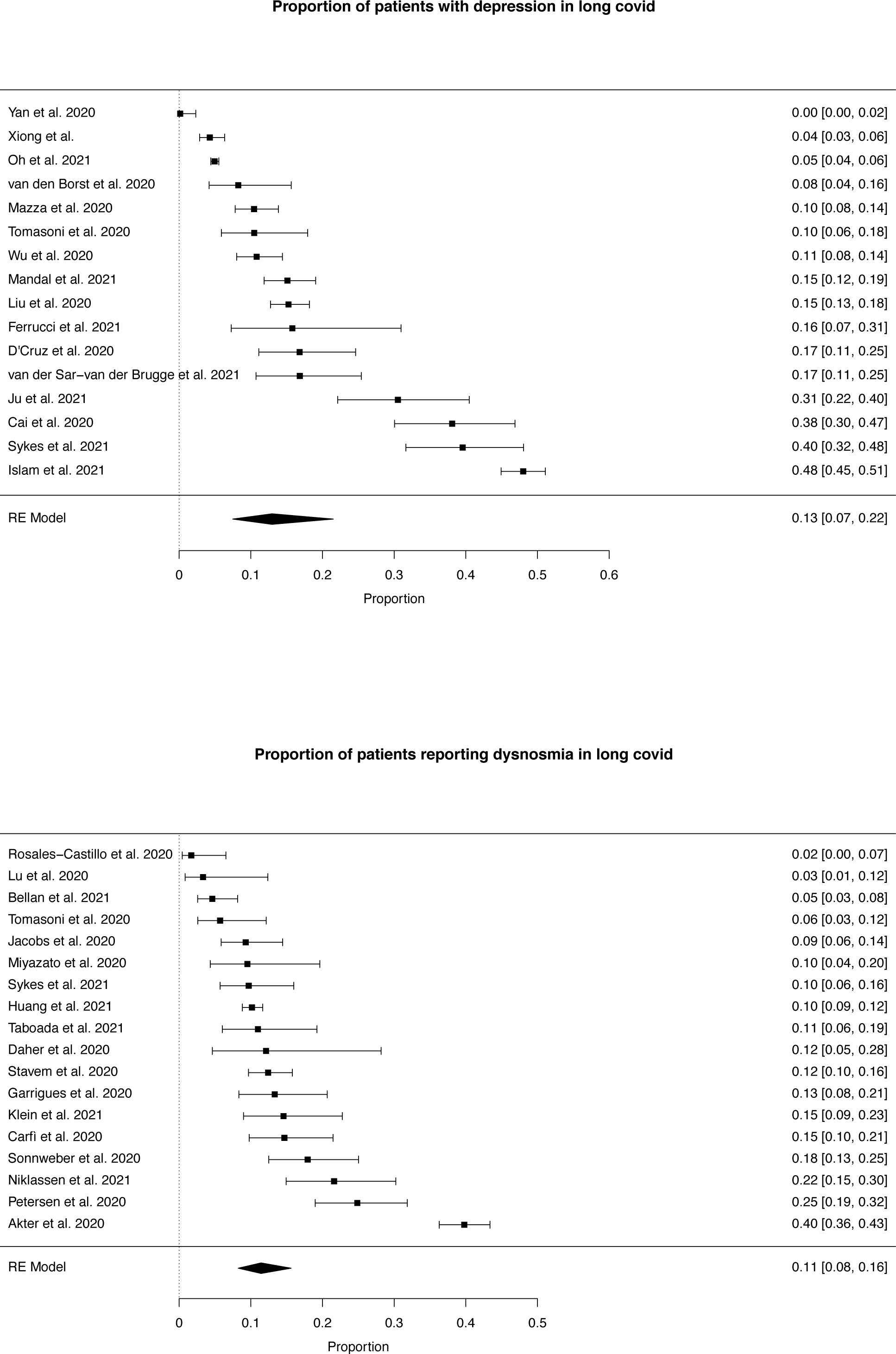
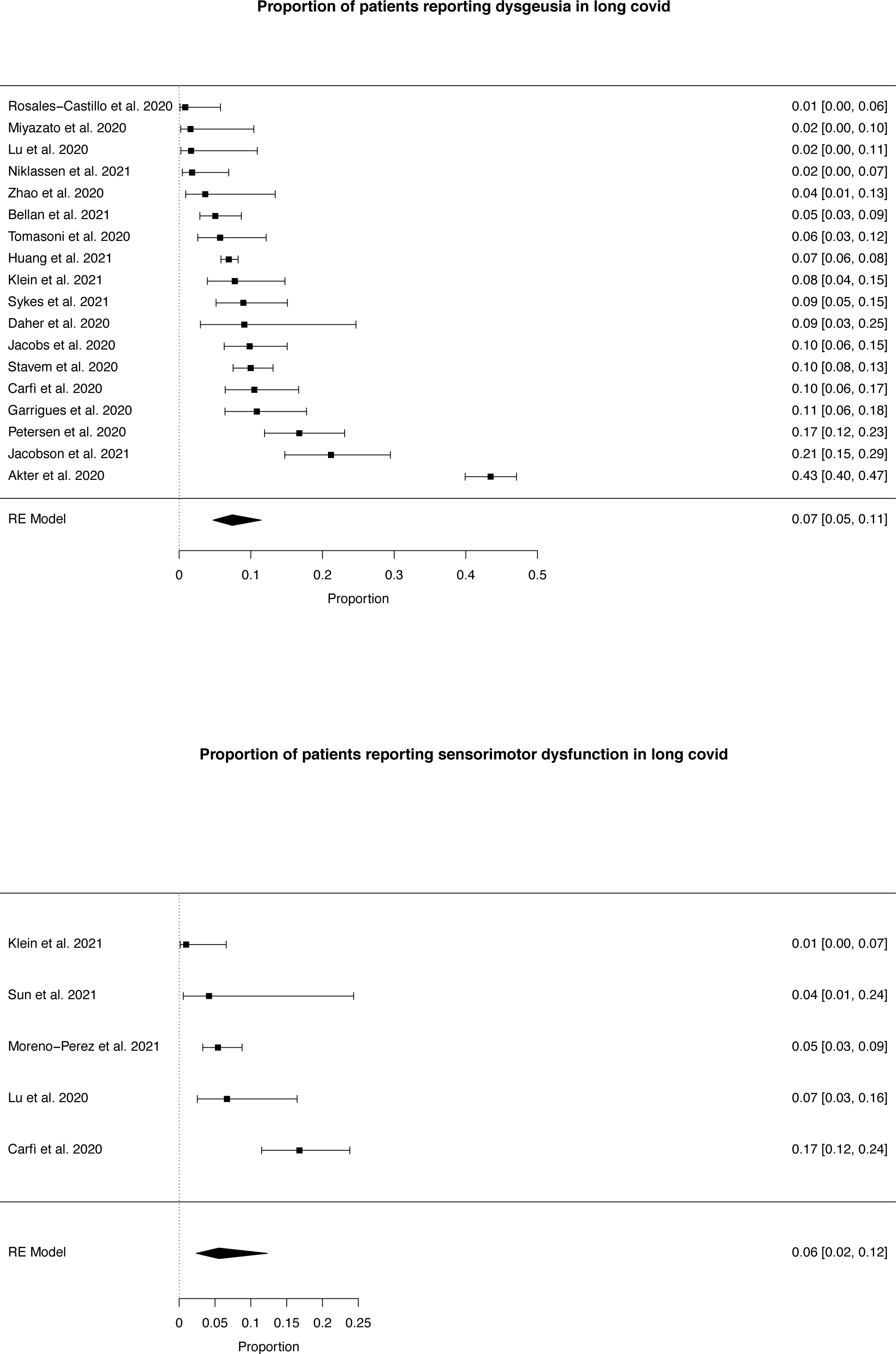
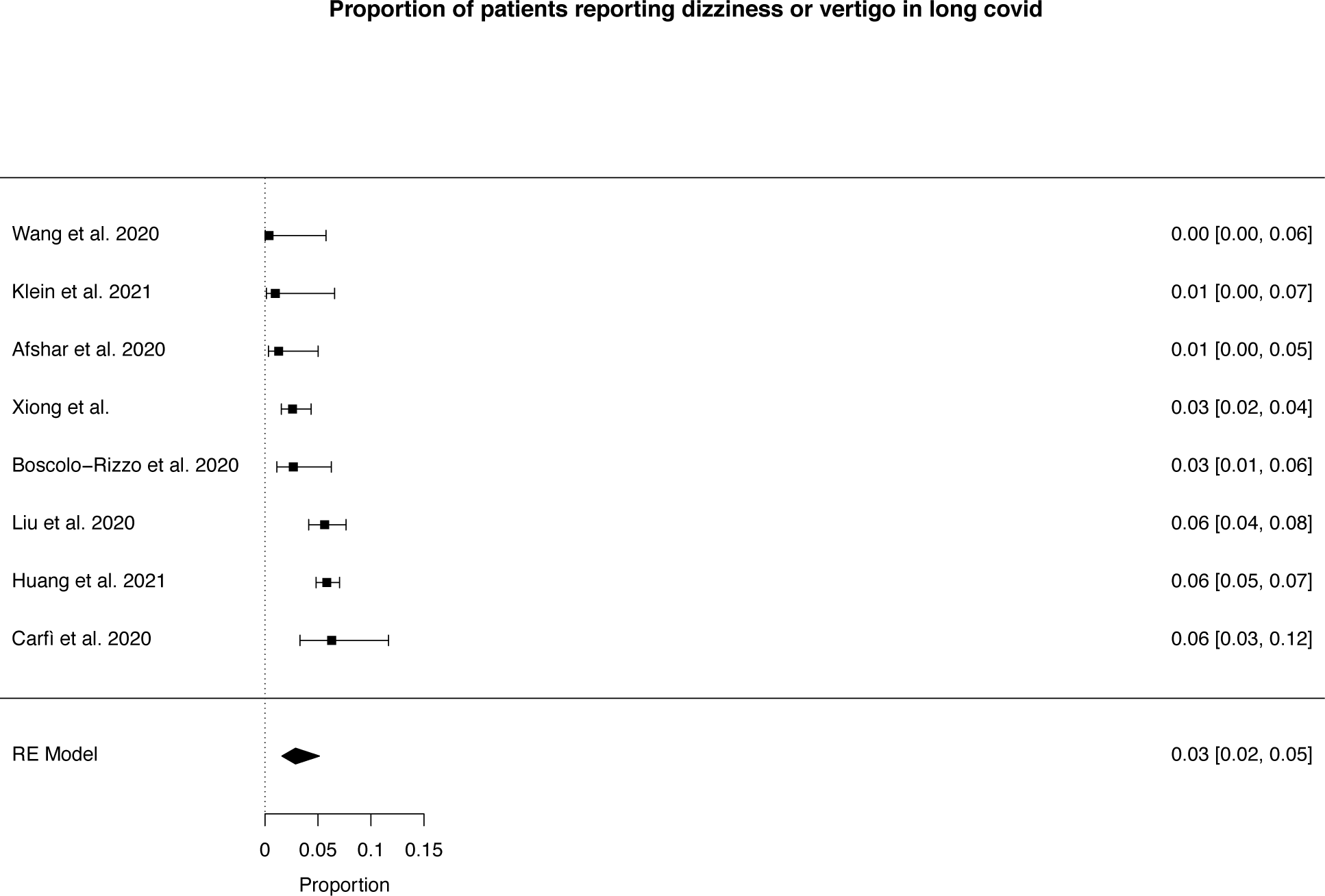
Forest plots for individual neuropsychiatric symptoms, all studies. Symptoms are plotted individually. The point prevalence for individual studies is presented with 95%confidence intervals on the right hand side of each plot. The pooled prevalence and 95% confidence interval for that symptom is shown at the bottom of each plot.

Only two studies reported symptoms in control groups, both drawn from healthy populations and neither formally matched to the respective COVID-19 groups. (36, 51) Each reported higher frequencies of sleep disorder, fatigue, dizziness, depression, anxiety, and/or psychosis in COVID-19 survivors compared to controls (Table S8).

### Secondary analyses

With the exception of anxiety, which was reported more frequently in non-hospitalised samples, there was no evidence of a differential prevalence of any symptom among hospitalised versus non-hospitalised samples (eight symptoms eligible to be tested), nor among patients admitted to ICU/having WHO ‘critical’ or ‘severe’ illness versus those not requiring ICU (six symptoms). Similarly there was no evidence of difference among patients surveyed <12 weeks versus 12+ weeks post-discharge (eight symptoms eligible to be tested), nor the same time-points post-symptom onset/PCR test (four symptoms, Figure 4, Table 3). Scatterplots of symptom prevalence reported by individual studies, plotted against duration since COVID-19, are presented in Figures S2 and S3.

**Figure 4.**
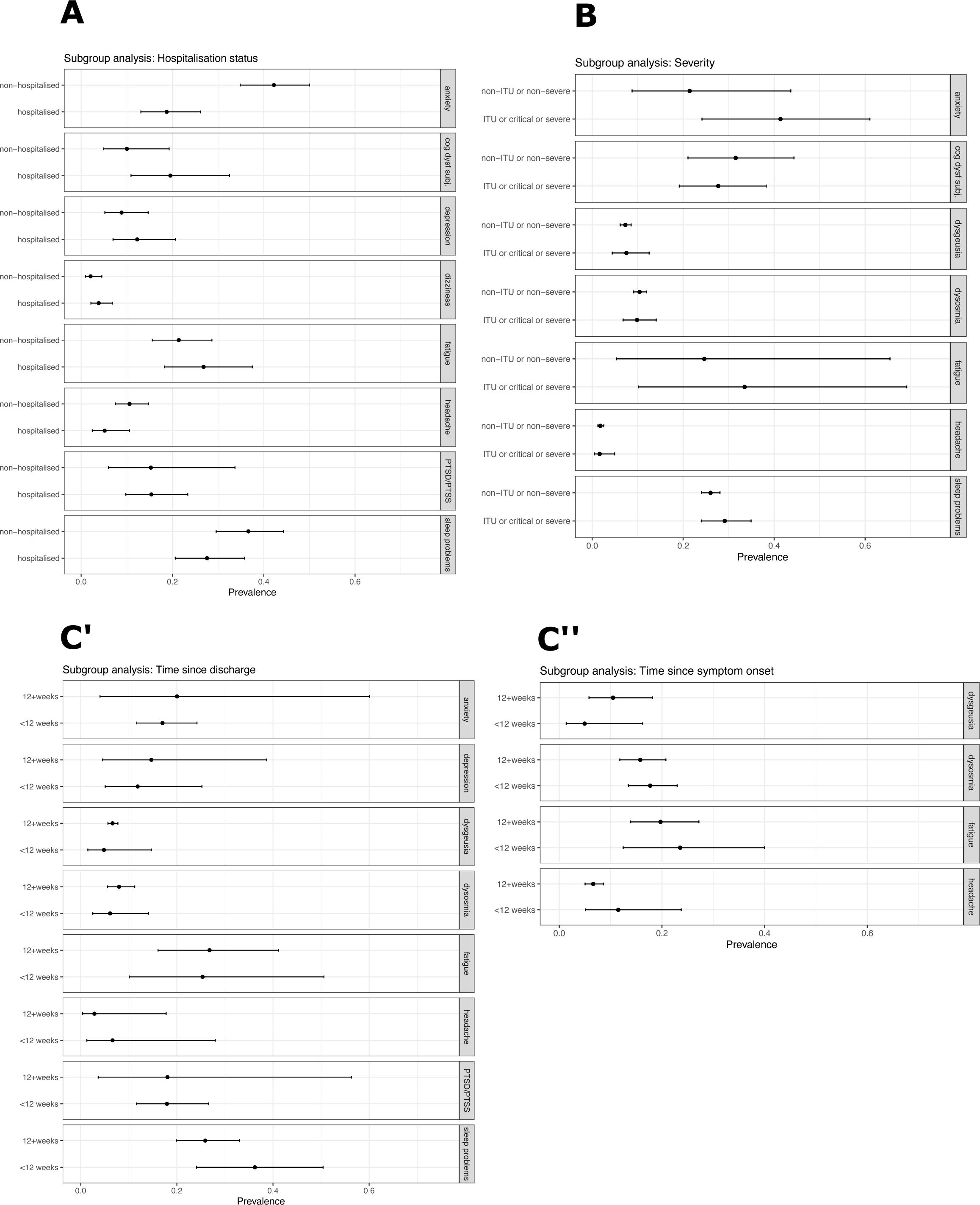
Pooled symptom prevalence by subgroups. Four subgroup analyses are shown (major panels **A-C’’**). Within each analysis symptoms which were eligible for analysis are plotted individually (identified in the right-hand tab on each minor panel). **A:** Comparison of pooled prevalence for studies reporting non-hospitalised versus hospitalised samples. **B:** Comparison of studies reporting patients who had non-ITU/non-severe versus ITU/critical/severe COVID-19. **C’:** Comparison of studies reporting duration of follow-up shorter than 12 weeks post-hospital discharge, versus those reporting longer follow- up. **C’’:** Comparison of studies reporting duration of follow-up shorter than 12 weeks since onset of COVID-19 symptoms, versus those reporting longer follow-up.

## DISCUSSION

In this systematic review and meta-analysis, we found that neuropsychiatric symptoms are common and persistent after COVID-19. Sleep disorders and fatigue appear to be especially prevalent and may be experienced by as many as one in four patients. Anxiety and post-traumatic stress symptoms also seem particularly common, and cognitive impairment is often objectively detectable. Sensorimotor disturbances and dizziness or vertigo are less common but present in a non-negligible proportion of patients. The prevalence of these symptoms appears to be relatively stable across different points in the first six months, between hospitalised and community samples, and among hospitalised patients regardless of COVID-19 severity. There are knowledge gaps in the neuropsychiatric consequences of COVID-19 in patients who did not require hospital admission, the impact of ethnicity, and the course and frequency of symptoms in the longer term.

These findings should be interpreted cautiously. Three in five studies in this review reported symptoms within the NICE guideline-suggested threshold of twelve weeks for the post-acute phase. (9) Relatively few eligible community-based or ICU-admitted samples reported our outcomes of interest, making rounded conclusions about the impact of COVID-19 severity difficult to draw. If, in due course, significant symptomatic differences emerge from data comparing hospitalised and non-hospitalised patients, then there could be a case that the term “Long COVID” is best reserved for patients who weren’t hospitalised - or that a subspecifier could be useful to denote severity of initial respiratory and/or other symptoms. Non-hospitalised patients were in the minority in this review (with only 15·7% confirmed as such), reflecting the early research focus on hospitalised patients. However, non-hospitalised patients were the majority (91·6%) in a recent large patient-led survey. In our view patient perspectives on terminology for this initially patient-driven disorder should be considered equally alongside those of clinicians and researchers. (15) Most studies tended to report outcomes based on patient self-report rather than structured clinical assessments. Meanwhile the lack of active control groups meant that the specific contribution of COVID-19 to such neuropsychiatric symptoms remains unknown. It is possible that the breadth and frequency of these symptoms represent the natural trajectory of recovery from a serious viral illness. The extent to which neuropsychiatric symptoms were new-onset, versus relapse of an existing condition, was not reported. Nor did we formally appraise the eligibility of 15 studies in which there was no English-language article available.

Our pooled data however imply frequent neuropsychiatric morbidity among COVID-19 survivors in the post-acute phase. These observations echo a recent large study associating COVID-19 with an increased risk of neuropsychiatric clinical diagnoses in the first six months, including first-onset insomnia, mood, anxiety, or psychotic disorders, and dementia. (14) The same study found a higher risk of such disorders after COVID-19 compared to other respiratory tract illnesses, indicating that at least some of the apparent neuropsychiatric burden may be COVID-19-specific. Notably the trajectory of accrual of new psychiatric diagnoses flattened only slightly in the first six months, supporting the hypothesis that neuropsychiatric symptoms persist within this timeframe. Our data also broadly support the aforementioned patient-led survey, of 3762 mainly non-hospitalised COVID-19 patients, (15) in which fatigue, self-reported cognitive dysfunction, and other neuropsychiatric symptoms (e.g. dizziness and sensorimotor symptoms) were highly prevalent. Owing to the self-selected nature of that study population - which was recruited mostly via Long COVID support groups and similar organisations - their data would be ineligible to contribute to generalisable estimates of community prevalence in the current meta-analysis; a characteristic which illustrates the difficulty of finding generalisable community-based samples. (79) The extent to which neuropsychiatric symptom burden impacts upon clinical services remains to be seen, although structuring Long COVID services to be proactive in case identification and treatment seems sensible. It remains possible that the commonest symptoms, such as (in descending order of frequency) insomnia, fatigue, anxiety, cognitive impairment, post-traumatic stress, and depression may respond to physical and/or psychological treatments as they do in other conditions. (80–84) In some instances, persisting symptoms after COVID-19 may reflect initial direct tissue injury mechanisms (e.g. inflammation) overlapping with other or additional mechanisms (e.g. cognitive) as can be seen in other complex disorders arising after illness like chronic pain. Multidisciplinary approaches are often appropriate for such disorders (85) including combinations of neurology, neurorehabilitation, neuropsychiatry, physiotherapy, occupational therapy and psychological input. Such approaches should be incorporated into the planning for ‘Long Covid’ services.

Our results identify areas for further research. Controlled studies are required to separate out the neuropsychiatric consequences of viral illness in general from those potentially specific to COVID-19 in particular. The impact of ethnicity and of COVID-19 severity remain to be clarified. Classical epidemiological approaches may be required to generate representative community-based samples, and longer-term follow-up is required. Emerging prospective, longitudinal, and multicentre studies will probe the characteristics and aetiology of persistent neuropsychiatric symptoms in patients with COVID-19. (86) Future trials meanwhile may examine treatments known to be effective in treating neuropsychiatric symptoms in other populations.

## Conclusion

Neuropsychiatric symptoms are common and persistent after recovery from COVID-19. Sleep problems and fatigue predominate and appear to affect roughly one quarter of survivors. Cognitive impairment, anxiety, post-traumatic symptoms, and depression are also common in the first six months. There is as yet little evidence that these persisting symptoms relate to the severity of, or duration since, initial infection. Although more research is needed, these early signals suggest a high burden of neuropsychiatric symptoms among COVID-19 survivors. Multidisciplinary services should be resourced accordingly in the post-COVID era.

## Supporting information

Tables S1-S8 (except S5) and Suppl Methods.

## Data Availability

Our full R code, and data, can be made available on reasonable request.

## CONTRIBUTORS

JB, TRN, JR, MB, and AGR conceived the review. JB and ERR led the project. JB and AGR ran the searches. All authors bar CW, KJ, HH, MB, TP, IK, BM, and TRN extracted data. CW and KJ did the statistical analysis. AGR, JB, KJ, SC, and EB wrote the first draft of the report. All authors had the opportunity to comment and all approved the final version. HH, MB, TP, IK, BM, TRN, JPR, and AGR provided executive oversight and/or direct supervision. JB and AGR verified the data. All authors had full access to all the data in the study and had final responsibility for the decision to submit for publication.

## DATA SHARING

Our full R code, and data, can be made available on reasonable request.

## ACKNOWLEDGEMENTS

JPR is supported by the Wellcome Trust (102186/B/13/Z). IK is funded through the NIHR (Oxford Health Biomedical Research Facility, Development and Skills Enhancement Award) and the Medical Research Council (Dementias Platform UK and Deep and Frequent Phenotyping study project grants). HH is funded by the German Research Foundation (DFG, Grant: HO 1286/16-1).

---

**Figure S1.**
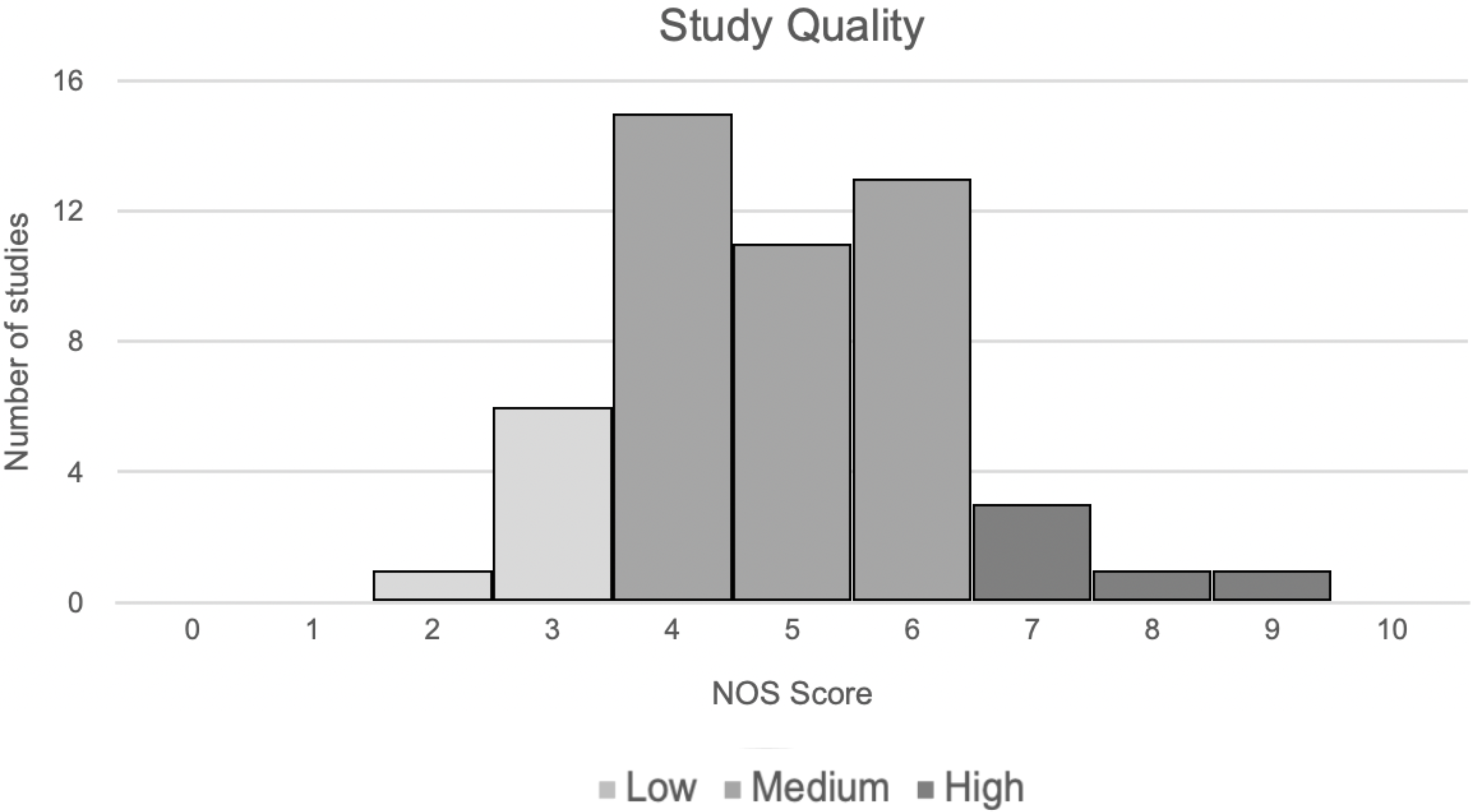
Histogram of study quality scores. The final Newcastle-Ottawa Scale (NOS) score for each included study is plotted in a histogram (total n=51 studies).

**Figure S2.**
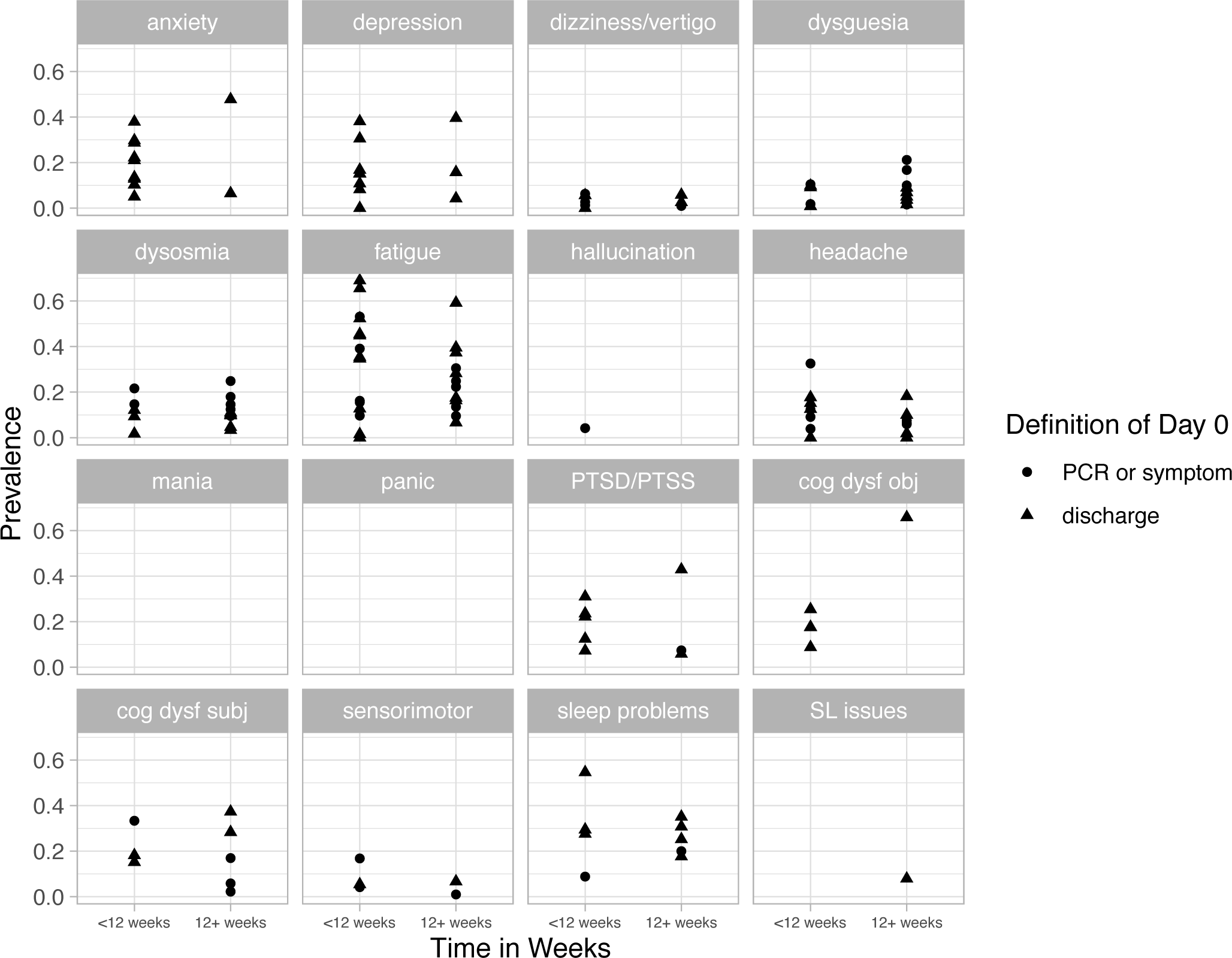
Scatterplots of symptom prevalence over time (dichotomised at 12 weeks). Each individual symptom is displayed in a minor panel. The point prevalence of that symptom in each individual study is represented by an individual data point. Studies are grouped according to their duration of follow-up (<12 versus 12+ weeks). The anchor point for follow-up duration (since the onset of symptoms/PCR testing, or since discharge) is indicated by the shape of each data point (see legend). In general there is little evidence of differential prevalence according to duration of follow-up.

**Figure S3.**
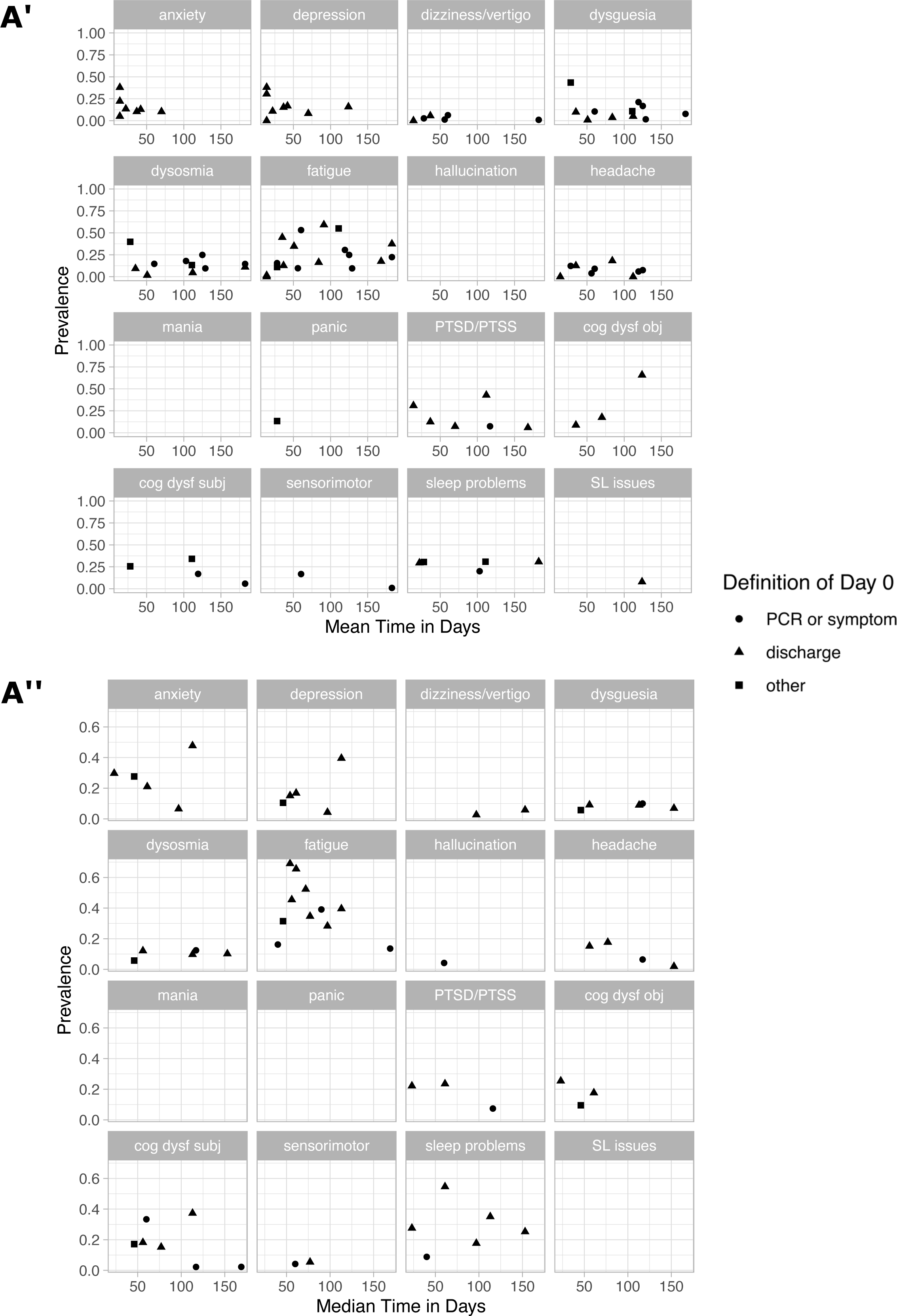
Scatterplots of symptom prevalence over time (continuous). Qualitative illustration of point prevalence estimates of individual symptoms (each minor panel) in individual studies (each data point). The anchor for follow-up duration is indicated by the shape of each point. Due to the relatively small number of studies for most symptoms, a quantitative analysis (e.g. a regression line) has not been conducted. **A’:** Scatterplots of studies reporting *mean* duration of follow-up. **A’’:** Scatterplots of studies reporting *median* duration of follow-up. In general there is little qualitative evidence of a marked change in the reported point prevalence of symptoms with increasing duration since COVID-19.

## LIST OF TABLES

Table S1. List of author contributions.

Table S2. Full list of data fields extracted from eligible studies.

Table S3. All secondary analyses conducted on each symptom.

Table S4. Summary of included studies.

Table S5. Rationale for excluding studies

Table S6. Ranking of study quality.

Table S7. Meta-analysis sensitivity analysis.

Table S8. Comparison of studies including control group.

