## Supplementary material for "Persistent neuropsychiatric symptoms after COVID-19: a systematic review and meta-analysis": Tables S1-S8 (except S5) and Suppl Methods.

Table S1. List of author contributions.

| Author | Contribution(s) |
| --- | --- |
| *All authors* | - Made a substantial intellectual contribution to the study - Approved the final manuscript |
| Mr James Badenoch | - Conceptualised study - Led and coordinated study - Screened studies for eligibility - Compared to other systematic reviews - Determined meta analysis eligibility - Data extraction methods, extracted data - Checked data extraction - Assisted in cleaning data for analysis - Supported with meta-analysis methods - Assisted in drafting manuscript sections - Arranged funding/COI statements |
| Dr Emma Rachel Rengasamy | - Screened studies for eligibility - Extracted data - Checked data extraction - Supervised quality assessment - Arbitrated quality assessment - Made supplementary table with complete list of studies - Checked adherence to PRISMA guidelines - Adapted to house style |
| Dr Cameron Watson | - Data extraction methods - Supported with meta-analysis methods - Conducted meta-analyses |
| Katrin Jansen | - Supported with meta-analysis methods - Conducted meta-analyses - Assisted with creating tables of results - Created tables of descriptive statistics |
| Miss Stuti Chakraborty | - Screened studies for eligibility - Extracted data - Made supplementary table with complete list of studies - Assisted in writing methods |
| Miss Ritika Dilip Sundaram | - Extracted data - Assisted in cleaning data for analysis - Conducted quality assessment - Made PRISMA flowchart - Created table of excluded studies |
| Mr Danish Hafeez | - Screened studies for eligibility - Extracted data - Data extraction methods- ethnicity - Assisted in cleaning data for analysis - Sorted references |
| Miss Ella Burchill | - Extracted data - Conducted quality assessment - Assisted in writing introduction |
| Mr Aman Saini | - Extracted data - Conducted quality assessment - Made supplementary table with complete list of studies - Created table of extracted data fields |
| Miss Lucretia Thomas | - Screened studies for eligibility - Extracted data - Conducted quality assessment - Made supplementary table with complete list of studies |
| Dr Benjamin Cross | - Screened studies for eligibility - Extracted data - Conducted quality assessment |
| Ms Camille Kaitlyn Hunt | - Screened studies for eligibility - Conducted quality assessment - Made table with characteristics of included studies |
| Miss Isabella Conti | - Extracted data - Conducted quality assessment |
| Ms. Sylvia Ralovska | - Checked extracted data |
| Dr Zain Hussain | - Conducted quality assessment |
| Dr Matthew Butler | - Conceptualised study - Arbitrated quality assessment - Assisted in writing manuscript - Provided senior review of manuscript |
| Dr Thomas Pollak | - Provided senior review of manuscript |
| Dr Ivan Koychev | - Provided senior review of manuscript |
| Dr Benedict Daniel Michael | - Provided senior review of manuscript |
| Prof Dr Heinz Holling | - Provided senior review of manuscript - Supervised application of meta-analytic methods |
| Dr Timothy R Nicholson | - Conceptualised study - Provided senior review of manuscript - Provided senior leadership and advice throughout |
| Dr Jonathan P Rogers | - Conceptualised study - Data extraction methods - Eligibility of studies for data extraction - Provided senior review of manuscript - Provided senior leadership and advice throughout |
| Dr Alasdair G Rooney | - Conceptualised study - Screened studies for eligibility - Consulted on screening studies - Data extraction methods - Extracted data - Checked data extraction - Supported with meta-analysis methods - Created figure of excluded studies - Created tables of descriptive statistics - Wrote first draft of manuscript - Provided senior leadership and advice throughout |

Table S2. Full list of data fields extracted from eligible studies.

Data field

First author of study, year

Study populations (specify individually reported subgroups on separate rows)

Treatment setting

Study design

Eligibility for meta-analysis (reason if not)

Eligibility for subgroup analysis

Need to contact authors with reason

Details on data extractions per study

Study country/countries of origin

Data collection start date in study

N infected at long-covid timepoint

N infected at long-covid timepoint PCR confirmed

N infected at long-covid timepoint not PCR confirmed

Number of patients from community

Number of patients post-discharge

Number of patients reported as 'WHO severe COVID-19'

Number of patients reported as 'WHO critical COVID-19'

Number of patients admitted to ICU/ITU/intensive care unit

Population age (mean)

Population age (standard deviation)

Population age (median)

Population age (IQR Q1)

Population age (IQR Q3)

Sex (n male)

Ethnicity descriptors stated by paper

Ethnicity – White

Ethnicity – Asian

Ethnicity – Black

Ethnicity – Mixed/Multiple Ethnic Groups

Ethnicity – Hispanic (South American)

Ethnicity – Other

Long COVID period defined from (post discharge, post PCR test etc.)

Mean long COVID duration (days)

Standard deviation long-COVID duration (days)

Median long-COVID duration (days)

IQR Q1 long-COVID duration (days)

IQR Q3 long-COVID duration (days)

Long-COVID duration categorised (< or > 12 weeks)

Definition of control group, if used

N control group

Control group matched variable(s)

Then we recorded the variables below for each symptom of interest (see footnote).

n infected reported with *x**

How *x* was reported in paper*

Diagnostic method*

Rating scale name with diagnostic threshold (if used) *

Mean rating scale score for infected participants (if used) *

Standard deviation rating scale score for infected participants (if used) *

Median rating scale score for infected participants (if used) *

IQR (Q1) rating scale score for infected participants (if used) *

IQR (Q3) rating scale score for infected participants (if used) *

Mean rating scale score for control participants (if used) *

Standard deviation rating scale score for control participants (if used) *

Median rating scale score for control participants (if used) *

IQR (Q1) rating scale score for control participants (if used) *

IQR (Q3) rating scale score for control participants (if used) *

** Data collected for each of the following neuropsychiatric symptoms: anxiety; depression; mania; panic; hallucinations; sleep changes; objectively reported cognitive dysfunction; subjectively reported cognitive dysfunction; sensorimotor symptoms (such as paraesthesia, numbness, or weakness of specific body parts), dizziness and vertigo; headache; changes in speech or language; and changes in taste or smell; fatigue; and post-traumatic stress disorder or symptoms.*

Table S3. All secondary analyses conducted on each symptom.

| SYMPTOM | Included in main meta-analysis? | *If 'N', state reason* | Included in "hospitalised vs non-hospitalised" analysis? | *If 'N',*  *state reason* | Included in "severity" analysis? | *If 'N',*  *state reason* | Included in "time" analysis 1: post-discharge <12/ 12+ weeks? | *If 'N',*  *state reason* | Included in "time" analysis 2: post-symptom/PCR onset <12/ 12+ weeks? | *If 'N',*  *state reason* |
| --- | --- | --- | --- | --- | --- | --- | --- | --- | --- | --- |
| Obj cog dysfunction | Y | - | N | <2 non-hosp studies | N | <5 pops | N | <5 pops | N | <5 pops |
| Subj cog dysfunction | Y | - | Y | - | Y | - | N | <5 pops | N | <5 pops |
| Sensorimotor | Y | - | N | <5 populations | N | <5 pops | N | <5 pops | N | <5 pops |
| Dizziness/ vertigo | Y | - | Y | - | N | <5 pops | N | <5 pops | N | <5 pops |
| Sleep | Y | - | Y | - | Y | - | Y | - | N | <5 pops |
| depression | Y | - | Y | - | N | <5 pops | Y | - | N | <5 pops |
| Anxiety | Y | - | Y | - | Y | - | Y | - | N | <5 pops |
| PTSD | Y | - | Y | - | N | <5 pops | Y | - | N | <5 pops |
| Dysguesia | Y | - | N | <2 non-hosp studies | Y | - | Y | - | Y | - |
| Dysosmia | Y | - | N | <2 non-hosp studies | Y | - | Y | - | Y | - |
| Fatigue | Y | - | Y | - | Y | - | Y | - | Y | - |
| Headache | Y | - | Y | - | Y | - | Y | - | Y | - |
| Speech and language | N | <3 studies | - | - | - | - | - | - | - | - |
| Panic | N | <3 studies | - | - | - | - | - | - | - | - |
| Mania | N | <3 studies | - | - | - | - | - | - | - | - |
| Hallucination | N | <3 studies | - | - | - | - | - | - | - | - |
| To be eligible for the main meta-analysis, a symptom had to have been reported by a minimum of three eligible studies. | | | | | | | | | | |
| Eligibility for subgroup analysis required both of the following: i) 2+ studies in each subgroup, and ii) 5+ study populations spread across the two sub-groups.  One study could report hospitalised and non-hospitalised prevalences separately, i.e. contribute two “populations”. | | | | | | | | | | |

Table S4. Summary of included studies.

Outcomes refer to all COVID-19 infected patients. Mean values given, where median is used, illustrated by *, IQR/Range specified.

| Ref | Study | Setting | Population | Design | N (number of patients with COVID-19) | Follow-up duration, mean (sd) | Mean age (sd) | Male (%), Female (%) | Prevalence outcomes (%) | Other outcomes reported, mean (sd) |
| --- | --- | --- | --- | --- | --- | --- | --- | --- | --- | --- |
| (1) | Lorenzo et al· 2020 | Italy | Mixed | Cohort | 185 | Post-discharge 23* (IQR 20-29) | 57* (IQR 48-67)  ‘ | Male 123 (66·5), female 62 (33·5) | Objective cognitive dysfunction 47 (25·4)  Sleep change  51 (27·6)  Anxiety 55 (29·7)  PTSD 41 (22·2) | - |
| (2) | Ju et al· 2020 | China | Hospitalised | Cohort | 95 | Post-discharge  14 (-) | 39* (IQR 30-47) | Male 51 (53·6)  Female 44  (46·3) | Depression 29 (30·5)  Anxiety (37·8) | PHQ-9  2* (Range 0-7)  GAD-7  2* (Range 0-6) |
| (3) | Simani et al· 2021 | Iran | Hospitalised | Cohort | 120 | Post-discharge 168 (-) | 54·6 (16·9) | Male 80 (66·7)  Female 40 (33·3) | PTSD 7 (5·8)  Fatigue 21 (17·5) | PCL-5  9·3 (10·8)  (-) |
| (4) | Niklassen et al· 2021 | Italy/Germany | Mixed | Cohort | 111 | Post-PCR test (-) | 44·5 (15) | Male 59 (53·2)  Female 52 (46·8) | Dysguesia 2 (1·8)  Dysnosmia 24 (21·6) | TDI 12·5 (1·5) |
| (5) | Tomasoni et al· 2020 | Italy | Hospitalised | Cross−sectional | 105 | Virological clearance 46* (IQR 43-48) | 55* (IQR 43-65) | Male 77 (73·3)  Female 28 (26·7) | Memory disorder 18 (17·1)  Depression 11 (10·5)  Anxiety 29 (27·6)  Dysguesia 6 (5·7)  Dysnosmia 6 (5·7)  Fatigue 33 (31·4) |  |
| (6) | Klein et al· 2021 | Israel | Non-hospitalised | Cohort | 103 | Post-PCR test  182·5 (-) | 35 (12) | Male 64 (62·1)  Female 39 (37·8) | Memory disorder 6 (5·8)  Paraesthesia 1 (0·9)  Dizziness/ Vertigo 1 (0·9)  Dysguesia 8 (7·7)  Dysnosmia 15 (14·5)  Fatigue 23 (22·3) |  |
| (7) | Taboada et al· 2021 | Spain | Hospitalised | Cohort | 91 | Post-discharge 182·5 (-) | 65·5 (10·4) | Male 59 (64·8)  Female 32 (35·1) | Sleep change 28 (30·7)  Dysnosmia 10 (10·9)  Fatigue 34 (37·3)  Anxiety/ Depression 42 (46·1) |  |
| (8) | Bellan et al· 2021 | Italy | Hospitalised | Cohort | 238 | Post-discharge 112 (-)  PCR confirmed 238  PCR not  confirmed 6  ICU Admission 28 | 61* (IQR 50-71) | Male 142 (59·6)  Female (40·3) | PTSD 102 (42·8)  Dysguesia 12 (5·04)  Dysnosmia 11 (4·6) | IES-R 0 (0) |
| (9) | Oh et al· 2021 | South Korea | Mixed | Cohort | 5879  Control group n = 93,683 | Other (-) | (-) | (-) | Depression 291 (4·9)  Hallucination 10 (0·2) |  |
| (10) | Miyazato et al· 2020 | Japan | Hospitalised | Cohort | 63 | Post-symptom onset 129 (21) | 48·1 (18·5) | Male 42 (66·7)  Female 21 (33·3) | Dysgeusia 1 (1·5)  Dysnosmia 6 (9·5)  Fatigue 6 (9·5) |  |
| (11) | Liang et al· 2020 | China | Hospitalised | Cohort | 76 | Post-discharge 91 (-) | 41·3 (13·8) | Male 21 (27·6)  Female 55 (72·3) | Fatigue 45 (59·2) |  |
| (12) | Sonnweber et al· 2020 | Austria | Mixed | Cohort | 145  Community 36  Hospitalised 109 | Post-PCR test 103 (21) | 57 (14) | Male 82 (56·6)  Female 63 (43·4 ) | Sleep change 29 (20)  Dysnosmia 26 (17·9) |  |
| (13) | Garrigues et al· 2020 | France | Hospitalised | Cross−sectional | 120 | Other 110·9 (11·1) | 63·2 (15·7) | Male 75 (62·5)  Female 45 (37·5) | Attention 32 (26·6)  Memory 41 (34·1)  Sleep change 37 (30·8)  Dysguesia 13 (10·8)  Dysnosmia 16 (13·3)  Fatigue 66 (55) |  |
| (14) | Afshar et al· 2020 | USA | Mixed | Cohort | Baseline:594  Week 4: 334  Week 6: 267  Week 8: 155 | (-) (Wk0,4,6,8) | 31·3 (5·1) | Male 0  Female 594 (100) | Week 4: Headache 23 (6·9), Dizziness/fainting 13 (3·9), Fatigue 40 (12·0)  Week 6: Headache 21 (7·9), Dizziness/fainting 3 (1·1), Fatigue 26 (9·7)  Week 8: Headache 6 (3·9), Dizziness/fainting 12 (1·3), Fatigue 15 (9·7) |  |
| (15) | He et al· 2020 | China | Hospitalised | Cohort | 420 | Post-symptom onset | 56* (IQR 43-63·8) | Male 207 (49·3)  Female 213 (50·7) | Sleep change 37 (8·7)  Fatigue 68 (16·2) |  |
| (16) | Akter et al· 2020 | Bangladesh | Hospitalised | Cross−sectional | 734 | Virological clearance 28 (-) | (-) | Male 558 (76·0)  Female 176 (24·0) | Loss of concentration 188 (25·6)  Memory loss 141 (19·2)  Sleep change 224 (30·5)  Panic disorder 98 (13·4)  Dysguesia 319 (43·4)  Dysnosmia 292 (39·7)  Fatigue 81 (11·0) |  |
| (17) | Zhao et al· 2020 | China | Hospitalised | Cohort | 55 | Post-discharge  84 (-) | 47·7 (15·5) | Male 32  (58·2)  Female 23 (41·8) | Headache 10 (18)  Dysguesia 2 (3·6)  Fatigue 9 (16·3) |  |
| (18) | D’Cruz et al· 2020 | UK | Hospitalised (ICU only) | Cohort | 119 | Post-discharge  61* (IQR 51-67) | 58·7 (14·4) | Male 74 (62·1)  Female 45 (37·8) | Cognitive Impairment 21 (17·6)  Sleep change 65 (54·6)  Depression 20 (16·8)  Anxiety 25 (21)  PTSD/PTS 28 (23·5)  Fatigue 78 (65·5) | 6-Item Cognitive Impairment Test (>/=8)  PHQ-9 (>9)  GAD-7 (>9)  Trauma Screen Questionnaire (>/=6) |
| (19) | Sun et al· 2021 | USA | Mixed | Cohort | 24  Community 17  Hospitalised 7 | Post-symptom onset  60* (IQR 40·8-85) | 45·3 (12·7) | Male 6 (25)  Female 18 (75) | Memory/Cognition 8 (33·3)  Sensorimotor (double vision) 1 (4·2)  Hallucinations 1 (4·2) |  |
| (20) | Islam et al· 2021 | Bangladesh | Mixed | Cross-sectional | 1002  Community 794  Hospitalised 208 | - | 34·7 (13·9) | Male 580 (57·9)  Female 422 (42·1) | Depression 481 (48)  Fatigue/asthenia 115 (11·5) | PHQ-9 9·08 (6·4)  9* |
| (21) | Huang et al· 2021 | China | Hospitalised | Cohort | 1733 | Other  186 (-) | 57* (IQR 47-65) | Male 897 (51·8)  Female 836 (48·2) | Headache 33 (1·9)  Dizziness/vertigo 101 (5·8)  Sleep change 437 (25·2)  Dysguesia 120 (6·9)  Dysnosmia 176 (10·2) |  |
| (22) | Moreno-Perez et al· 2021 | Spain | Mixed | Cohort | 277  Community 95  Hospitalised 182 | Other  77* (IQR 72-85) | 56 (-) | Male 146 (52·7)  Female 131(47·2) | Subjective cognitive dysfunction 42 (15·1)  Sensorimotor (visual loss) 15 (5·4)    Headache 49 (17·7)  Fatigue 96 (34·6) |  |
| (23) | Halpin et al· 2020 | UK | Hospitalised | Cohort | 100 | Post-discharge 48 (10·3) | - | Male 54 (54)  Female 46 (46) | Concentration/moeory 23 (23),  SLT issues 20 (20),  Fatigue 64 (64),  PTSD 31 (31) |  |
| (24) | Xiong et al | China | Hospitalised | Cohort | 538 | Post-discharge 97* (IQR 95-102) | 52* (41-62) | Male 245 (45·5)  Female 293 (54·4) | Dizziness/vertigo 14 (2·6)  Sleep change 95 (17·3)  Depression 23 (4·3)  Anxiety 35 (6·5)  Fatigue 152 (28·3) |  |
| (25) | Rosales-Castillo et al· 2020 | Spain | Hospitalised | Cohort | 118 | Post discharge 50·8 (6) | 60·2 (15·1) | Male 66 (55·9)  Female 52 (44·1) | Dysgeusia 1 (0·8)    Dysnosmia 2 (1·7)  Fatigue 41 (34·7) |  |
| (26) | Jacobs et al· 2020 | USA | Hospitalised | Cohort | 183 | Post discharge 35 (5) | - | Male 112 (61·2)  Female 71 (38·8) | Objective cognitive dysfunction 16 (8·7)  Headache 23 (12·6)  Dysgeusia 18 (9·8)  Dysnosmia 17 (9·3)  Fatigue 82 (44·8) |  |
| (27) | Daher et al· 2020 | Germany | Hospitalised | Cohort | 33 | Post discharge 56* (23) | 64 (3) | Male 22 (66·7)  Female 11 (33·3) | Subjective cognitive dysfunction 6 (18·2)  Headache 5 (15·2)  Anxiety (-)  Depression (-)  Dysgeusia 3 (9·1)  Dysnosmia 4 (12·1)  Fatigue 15 (45·5) | GAD-7 4*(8) |
| (28) | Arnold et al· 2020 | UK | Hospitalised | Cohort | 110 | Post symptom onset 90* (17) | - | Male 68 (61·8)  Female 42 (38·1) | Fatigue 43 (39·1) |  |
| (29) | Stavem et al· 2020 | Norway | Community | Cross-sectional | 451 | Post symptom onset 117* (162) | 49·8 (15·2) | Male 198 (43·9)  Female 253 (56·1) | Subjective cognitive dysfunction 10 (2·2)  Headache 29 (6·4)  Dysgeusia 45 (10)  Dysnosmia 56 (12·4) |  |
| (30) | Petersen et al· 2020 | Faroe Islands | Community | Cohort | 173 | Post symptom onset 125 (17) | 39·9 (19·4) | Male 82 (47·4)  Female 91 (52·6) | Headache 13 (7·5)    Dysgeusia 29 (16·8)  Dysnosmia 43 (24·9)  Fatigue 43 (24·9) |  |
| (31) | Boscolo-Rizzolo· 2020 | Italy | Community | Cohort | 187 | Post PCR test 28 | - | Male 84 (44·9)  Female 103 (55·1) | Headache 19 (23·7)  Dizziness 3 (12·0)  Fatigue 29 (13·9) |  |
| (32) | Van den Borst et al· 2020 | The Netherlands | Mixed | Cohort | 97 | Post discharge 70 (11·9) | - | Male 66 (68)  Female (32) | Depression 8 (8·2)  Anxiety 10 (10·3)  PTSD 7 (7·2) | HADS-depression 15(3·5)  HADS-anxiety 18(4·8)  PCL-5 14·5(15·9) |
| (33) | Carvalho-Schneider et al· 2021 | France | Mixed | Cohort | 103 | Post symptom onset | - | Male 44 (42·7)  Female 59 (57·3) | Headache 54 (52·4) |  |
| (34) | Mandal et al· 2021 | UK | Hospitalised | Cross-sectional | 384 | Post discharge 54* (12) | 59·9 (16·1) | Male 238 (62)  Female 146 (38) | Depression 58 (15·1)  Fatigue 265 (69) |  |
| (35) | Mazza et al· 2020 | Italy | Hospitalised | Cohort | 402 | Other | 57·8 (13·3) | Male 265 (66)  Female 137 (34) | Sleep problems 147 (36·6)  Depression 42 (10·4)    Anxiety 144 (35·8)  PTSD 52 (12·9) | WHIIRS 7·3 (5)  BDI-13 3·3 (4·4)  STAI-state 38·2 (11·1)  PCL 5 14·5 (15·9) |
| (36) | Pellaud et al· 2020 | Switzerland | Hospitalised | Cohort | 196 | Post symptom onset 30 | - | Male 119 (60·7)  Female 77 (39·3) | (-) |  |
| (37) | Lu et al· 2020 | China | Hospitalised | Cohort | 60 | Post discharge | 44·1 (16) | Male 34 (56·7)  Female 26 (43·4) | Subjective cognitive dysfunction 17 (28·3)  Sensorimotor 4 (6·7)  Headache 6 (10)  Dysgeusia 1 (1·7)  Dysnosmia 2 (3·3)  Fatigue 4 (6·7) |  |
| (38) | Zhu et al· 2020 | China | Hospitalised | Cohort | 432 | Post discharge | - | Male 225 (52·1)  Female 207 (47·9) | Anxiety 124 (28·7)  Fatigue 153 (35·4) |  |
| (39) | Jacobson et al· 2021 | USA | Mixed | Cohort | 118 | Post PCR test 119·3 (33) | 43·3 (14·4) | Male 63 (53·4)  Female 55 (46·6) | Subjective cognitive dysfunction 20 (16·9)  Headache 7 (5·9)  Dysgeusia 25 (21·2)  Fatigue 36 (30·5) |  |
| (40) | Skyes et al· 2021 | UK | Hospitalised | Cohort | 134 | Post discharge 113* | 59·6 (14) | Male 88 (65·7)  Female 46 (34·3) | Subjective cognitive dysfunction 50 (37·3)  Sleep problems 47 (35)  Depression 53 (39·6)  Anxiety 64 (47·8)  Dysgeusia 12 (9)  Dysnosmia 13 (9·7)  Fatigue 53 (39·6) |  |
| (41) | Van der Sar-van der Brugge et al· 2021 | The Netherlands | Hospitalised | Cohort | 101 | Post discharge 42 | 66·4 (12·6) | Male 58 (57·4)  Female 43 (42·6) | Depression 17 (16·8)  Anxiety 13 (12·9) | HADS-depression 3*(5)  HADS-anxiety 4*(5) |
| (42) | Townsend et al· 2020 | Ireland | Mixed | Cohort | 128 | Other 72* (25) | 49·5 (15) | Male 59 (46·1)  Female 69 (53·9) | Fatigue 67 (52·3) | Chalder Fatigue Scale 15·8 (5·9) |
| (43) | Wu et al· 2020 | China | Hospitalised | Cross-sectional | 370 | Post discharge 22 | 50·5 (13·1) | Male 203 (54·9)  Female 167 (45·1) | Sleep problems 109 (29·5)  Depression 40 (10·8)  Anxiety 50 (13·5) |  |
| (44) | Yan et al· 2020 | China | Hospitalised | Cohort | 337 | Post discharge 14 | - | Male 154 (45·7)  Female 183 (54·3) | Anxiety 17 (5·0)  Fatigue 5 (1·5) |  |
| (45) | Cai et al· 2020 | China | Hospitalised | Cohort | 126 | Post discharge 14 | 45·7 (14) | Male 60 (47·6)  Female 66 (52·4) | Depression 48 (38·1)  Anxiety 28 (22·2)  PTSD 39 (30·9) | SDS 47·3 (13·1)  SAS 43·2 (10·2)  PTSD-SS 45·5 (18·9) |
| (46) | Liu et al· 2020 | China | Hospitalised | Cross-sectional | 675 | Post discharge 36·7 | - | Male 317 (47)  Female 358 (53) | Dizziness/vertigo 38 (5·6)  Depression 103 (15·2)  Anxiety 70 (10·3)  PTSD 84 (12·4)  Fatigue 86 (12·7) | PHQ-9 5*(3-8)  GAD-7 4*(2-6)  PCL-5 12*(4-16) |
| (47) | Wang et al· 2020 | China | Hospitalised | Cohort | 131 | Post discharge 14 | - | Male 59 (45)  Female 72 (55) | Headache 5 (3·8)    Dizziness/vertigo (0)  Fatigue 7 (5·3) |  |
| (48) | Carfì et al·2020 | Italy | Hospitalised | Cohort | 143 | Post symptom onset 60·3 (30·6) | 56·5 (14·6) | Male 90 (62·9)  Female 53 (37·1) | Sensorimotor 24 (16·7)  Headache 13 (9·1)  Dizziness/vertigo 9 (6·3)  Dysgeusia 15 (10·5)  Dysnosmia 21 (14·7)  Fatigue 76 (53·1) |  |
| (49) | Einvik et al· 2021 | Norway | Mixed | Cohort | 583 | Post symptom onset 116* | 51·3 | Male 274 (47)  Female 309 (53) | PTSD 43 (7·4) |  |
| (50) | Logue et al· 2021 | USA | Mixed | Cohort | 177 | Post symptom onset 169* | 48 (15·2) | Male 76 (42·9)  Female 101 (57·1) | Subjective cognitive dysfunction 4 (2·2)  Fatigue 24 (13·6) |  |
| (51) | Ferrucci et al· 2021 | Italy | Hospitalised | Cohort | 38 | Post-discharge 124(34·2) | 53·5 (12·6) | Male 27 (71·1)  Female 11 (28·9) | Objective cognitive dysfunction 25 (65·7)  Speech and language issues 3 (7·9)  Depression 6 (15·8) | WLG 25·65 (5·23) |

PTSD = Post-traumatic stress disorder , WLG = Word List Generation-WLG, BDI-13= Beck's Depression Inventory (13 item, GAD-7 = Generalized anxiety disorder scale, PCL-5=  DSM-5 PTSD Checklist, SAS = Self-rating anxiety scale , SDS = Self-rating depression scale, PTSD-SS = Post-traumatic stress disorder self-rating scale , PHQ-9=  Patient Health Questionnaire-9, HADS= Hospital Anxiety and Depression Scale, , WHIRS = Women’s Health Initiative Insomnia Rating Scale, STAI-trait = State-Trait Anxiety Inventory , IES-R= The Impact of Event Scale-Revised, PROMIS= Patient-Reported Outcomes Measurement Information System , TDI = threshold, discrimination, and identification, SIT-12= Sniffin' Sticks 12 Identification set , ARTIQ= Acute Respiratory Tract Infection Questionnaire, CFQ = Cognitive failures questionnaire, SLT= Speech or language issues,

40.	Sykes DL, Holdsworth L, Jawad N, Gunasekera P, Morice AH, Crooks MG. Post-COVID-19 Symptom Burden: What is Long-COVID and How Should We Manage It? Lung. 2021 Feb 11;

Table S6. Ranking of study quality.

| Study quality | n | % |
| --- | --- | --- |
| Low | 7 | 13.7% |
| Medium | 39 | 76.5% |
| High | 5 | 9.8% |
| Total studies | 51 | 100.0% |

| Design | Number of studies | % |
| --- | --- | --- |
| Cohort | 43 | 84.3% |
| Cross−sectional | 8 | 15.7% |
| Grand Total | 51 | 100% |

Table S7. Meta-analysis sensitivity analysis.

| Symptom | N studies | N subjects | Pooled prevalence | 95% CI | I^2^ |
| --- | --- | --- | --- | --- | --- |
| Fatigue | 32 | 7501 | 0·271 | [0·203, 0·345] | 97·9 % |
| Dysosmia | 18 | 4738 | 0·123 | [0·085, 0·166] | 93·2 % |
| Dysgeusia | 18 | 4675 | 0·086 | [0·05, 0·129] | 94·71 % |
| Depression | 16 | 10402 | 0·155 | [0·092, 0·23] | 98·51 % |
| Headache | 15 | 4023 | 0·081 | [0·042, 0·13] | 95·16 % |
| Anxiety | 14 | 3716 | 0·206 | [0·141, 0·279] | 96·25 % |
| Sleep problems | 12 | 4991 | 0·280 | [0·217, 0·348] | 95·79 % |
| Subj· cog· dysf· | 12 | 2336 | 0·174 | [0·103, 0·258] | 95·37 % |
| PTSD/PTSS | 9 | 2545 | 0·170 | [0·099, 0·255] | 96·2 % |
| Dizziness | 8 | 3665 | 0·029 | [0·014, 0·049] | 85·77 % |
| Obj· cog· dysf· | 6 | 727 | 0·217 | [0·089, 0·381] | 95·68 % |
| Sensorimotor | 5 | 607 | 0·061 | [0·017, 0·124] | 82·69 % |

Table S8. Comparison of studies including control group.

| Ref | Symptom | COVID-19 patients | Control group |
| --- | --- | --- | --- |
| Oh et al· [31] |  | n=5879 | n=93863 |
|  | *Depression* | 291 (4·9%) | 977 (1·0%) |
|  | *Psychosis* | 10 (0·17%) | 34 (0·04%) |
|  |  | n=538 | n=184 |
| Xiong et al· [46] | *Depression* | 23 (4·3%) | 2 (1·1%) |
|  | *Dizziness* | 14 (2·6%) | 3 (1·6%) |
|  | *Sleep changes* | 95 (17·7%) | 9 (4·9%) |
|  | *Anxiety* | 35 (6·5%) | 3 (1·6%) |
|  | *Fatigue* | 152 (28·3%) | 17 (9·2%) |

Supplementary Methods. Search Strategy.

Search strategy on OVID (MEDLINE, EMBASE, and PsycINFO)

1.	(post-acute covid* or postacute covid* or post acute covid*).mp.

2.	(post covid* adj3 (illness* or syndrome* or symptom*)).mp.

3.	(prolonged adj3 covid*).mp.

4.	(persistent adj3 covid*).mp.

5.	(chronic adj3 covid*).mp.

6.	(long covid* or longcovid* or long-covid*).mp.

7.	((long haul* or longhaul* or long-haul*) adj3 covid*).mp.

8.	1 or 2 or 3 or 4 or 5 or 6 or 7 or 8

9.	Remove duplicates from 8

10. (chronic adj3 (complication* or infect* or symptom* or syndrome*)).mp.

11. (long haul* or long-haul* or longhaul*).mp.

12. ((long-term or long term or longterm) adj3 (complication* or consequence* or outcome*)).mp.

13. (Persistent adj3 (infecti* or symptom* or syndrome*)).mp.

14. (prolonged adj3 recovery).mp.

15. sequelae.mp.

16. rehabilitat*.mp.

17. 10 or 11 or 12 or 13 or 14 or 15 or 16

18. (Coronavirus or corona virus or coronavirinae or coronaviridae or betacoronavirus or Covid19 or Covid 19 or Covid-19 or nCoV or CoV 2 or CoV2 or CoV-2 or Sarscov2 or SARS-CoV-2 or 2019nCoV).mp.

19. 17 and 18

20. Remove duplicates from 19

21. 9 or 20

22. Remove duplicates from 21

23. Limit 22 to yr=”2020-Current”

Search strategy for CINAHL

(post-acute covid* or postacute covid* or post acute covid*).mp. or (post covid* adj3 or (illness* or syndrome* or symptom*)).mp. or (prolonged adj3 covid*).mp. or (persistent adj3 covid*).mp. or (chronic adj3 covid*).mp. or (long covid* or longcovid* or long-covid*).mp. or ((long haul* or longhaul* or long-haul*) adj3 covid*).mp.

OR

((chronic adj3 (complication* or infect* or symptom* or syndrome*)).mp. or (long haul* or long-haul* or longhaul*).mp. or ((long-term or long term or longterm) adj3 (complication* or consequence* or outcome*)).mp. or (Persistent adj3 (infecti* or symptom* or syndrome*)).mp. or (prolonged adj3 recovery).mp.

sequelae.mp.or rehabilitat*.mp.) AND (Coronavirus or corona virus or coronavirinae or coronaviridae or betacoronavirus or Covid19 or Covid 19 or Covid-19 or nCoV or CoV 2 or CoV2 or CoV-2 or Sarscov2 or SARS-CoV-2 or 2019nCoV).mp.
