## Supplementary material for "Persistent neuropsychiatric symptoms after COVID-19: a systematic review and meta-analysis": Table S5.

| Study | Year | 1. Is it original data? NO if e.g., any review, opinion piece, editorial | 2. Are the participants infected with SARS-CoV-2? e.g., see comment | 3. Are neuropsychiatric symptoms reported? See comment | 4. Was the duration post-COVID long enough? see comment | 5. Is the sample size 10 or more? | 6. If it's mechanistic or pre-clinical data - are symptoms reported? e.g., in comment | 7. Is the paper peer-reviewed? | 8. Is the paper in English language? | 9. Specific prevalences reported for individual symptoms | 10. Sample not sufficiently generalisable | 11. Ineligible study design | 12. Mark 'No' here if there's another reason for exclusion | Rationale |
|---|---|---|---|---|---|---|---|---|---|---|---|---|---|---|
| Lanham et al. 2021 | 2021 | | | | | | | | | No | | | | Specific prevalences not reported for individual symptoms |
| Liu et al. 2021 | 2021 | | | | | | | | | No | | | | Specific prevalences not reported for individual symptoms |
| Horn et al. 2020 | 2020 | | | | | | | | | | | | No | Could not find manuscript online |
| Barin et al. 2021 | 2021 | | | | | | | | | No | | | | Specific prevalences not reported for individual symptoms |
| Puchner et al. 2021 | 2021 | | | | | | | | | | No | | | Recruited patients referred to a rehabilitation programme for ongoing symptoms (enriched sample) |
| Machado et al. 2021 | 2021 | | | | | | | | | | No | | | Recruited members of two Facebook groups for COVID-19 subjects with persistent complaints (enriched sample) |
| Mandora et al. 2021 | 2021 | | | | | | | | | | No | | | Recruited patients admitted to a sub-acute care facility for ongoing symptoms (enriched sample) |
| Guedj et al. 2021 | 2021 | | | | | | | | | | No | | | Recruited patients with Long COVID (enriched sample) |
| Garg et al. 2021 | 2021 | | | | | | | | | | No | | | Reports data on patients with persisting fatigue only (symptom selection) |
| Cellai and O'Keefe, 2020 | 2020 | | | | | | | | | | No | | | Reports data on patients who requested ongoing follow-up for persisting symptoms (enriched sample) |
| Salmon-Ceron et al. 2021 | 2021 | | | | | | | | | | No | | | Reports data from a Long COVID outpatient clinic (enriched sample) |
| Goertz et al. 2020 | 2020 | | | | | | | | | | No | | | Reports data from members of two Facebook groups for coronavirus patients with persistent complaints (enriched sample) |
| Kandemirli et al. 2021 | 2021 | | | | | | | | | | No | | | Reports data on patients selected for having persistent COVID-19 related olfactory dysfunction (symptom selection) |
| Fjaeldstad. 2020 | 2020 | | | | | | | | | | No | | | Reports on patients selected for having smell loss for more than two weeks (symptom selection) |
| Meys et al. 2020 | 2020 | | | | | | | | | | No | | | Recruited patients from a Facebook support group for patients with persistent complaints after COVID-19 (enriched sample) |
| Chiesa-Estomba et al. 2020 | 2020 | | | | | | | | | | No | | | |
| Lovato et al. 2020 | 2020 | | | | | | | | | | No | | | Reports on patients selected for having olfactory and/or taste dysfunction (symptom selection) |
| Sofian et al. 2021 | 2021 | | | | | | | | | | No | | | Reports on patients selected for having prolonged symptoms (enriched sample) |
| Ladds et al. 2021 | 2021 | | | | | | | | | | No | | | Reports on patients recruited through online long COVID support groups and social media (enriched sample) |
| Liu et al. 2020 | 2020 | | | | | | | | | | | No | | RCT |
| Tang et al. 2021 | 2021 | | | | | | | | | | | No | | Uncontrolled interventional study |
| Neto et al. 2021 | 2021 | | | | No | | | | | | | | | Median 13-16 days post symptom onset |
| Kamal et al. 2020 | 2020 | | | | No | | | | | | | | | >20 days since negative PCR (virological clearance), mean/median however not stated. |
| Heywood et al. 2021 | 2021 | | | No | | | | | | | | | | No neuropsychiatric symptoms reported |
| Weerahandi et al. 2021 | 2021 | | | | | | | | | | | | No | Prevalences not reported |
| Roberts et al. 2021 | 2021 | | | No | | | | | | | | | | No neuropsychiatric symptoms reported |
| Caronna et al. 2020 | 2020 | | | No | | | | | | | | | | No persistent neuropsychiatric symptoms reported |
| Landsman et al. 2020 | 2020 | | | | | | | | No | | | | | Manuscript in Dutch |
| Buttner L. and Bauknecht H.C. and Fleckenstein | 2020 | | | | No | | | | | | | | | |
| Atalla E. and Zhang R. and Shehadeh F. and My | 2021 | | | | No | | | | | | | | | |
| Koutroumanidis M. and Gratwicke J. and Sharma | 2021 | No | | | | | | | | | | | | |
| Miller C. and O'Sullivan J. and Jeffrey J. and Pov | 2021 | | | No | | | | | | | | | | |
| Tenforde M.W. and Kim S.S. and Lindsell C.J. ar | 2020 | | | | No | | | | | | | | | |
| Li Z. and Zheng C. and Duan C. and Zhang Y. ar | 2020 | | | | No | | | | | | | | | |
| Woolford S.J. and D'Angelo S. and Curtis E.M. a | 2020 | | | No | | | | | | | | | | |
| Tenforde M.W. and Billig Rose E. and Lindsell C. | 2020 | | | | No | | | | | | | | | |
| Tian S. and Chang Z. and Wang Y. and Wu M. ar | 2020 | | | | No | | | | | | | | | |
| Sahanic S. and Sonnweber T. and Pizzini A. and | 2020 | | | | | | | | | | | | | |
| Lerum T.V. and Aalokken T.M. and Bronstad E. a | 2020 | | | | | | | | | | | | | |
| Juarez J. and Smith C. and Loo G. and Nestor N | 2020 | | | | | | | | | | | | No | |
| Paneroni M. and Simonelli C. and Saleri M. and | 2021 | | | No | No | | | | | | | | | |
| Dawson C. and Capewell R. and Ellis S. and Ma | 2020 | | | No | | | | | | | | | | |
| Fontaine C. | 2020 | No | | | | | | | | | | | | |
| Tuzun S. and Keles A. and Okutan D. and Yildira | 2021 | | | | No | | | | | | | | | |
| Miners S. and Kehoe P.G. and Love S. AO - Love | 2020 | No | | | | | | | | | | | | |
| Escalon M.X. and Herrera J. | 2020 | No | | | | | | | | | | | | |
| Yong, Shin Jie | 2021 | No | | | | | | | | | | | | |
| Pant P. and Joshi A. and Basnet B. and Shrestha | 2021 | | | | No | | | | | | | | | |
| Ladds E. and Rushforth A. and Wieringa S. and | 2020 | | | | | | | | | | | | No | |
| McCue C. and Cowan R. and Quasim T. and Pu | 2020 | | | No | | | | | | | | | | |
| Padman D. and Aya H.D. and Bhide A. and Low | 2020 | | | | | | | | | | | | No | |
| Messina A. and Andrea P. and Valeria A. and Ma | 2020 | | | | | | | | | | | | No | |
| Al-Hindawi A. and Sokhi J. and El-Hibri F. and Hi | 2020 | | | | | | | | | | | | No | |
| Burgers J. | 2020 | | | | | | | | | | | | No | |
| Peel M. | 2020 | | | | | | | | | | | | No | |
| Bruce S.S. and Kahan J. and Huq T. and Santilla | 2021 | | | No | | | | | | | | | | |
| Marcus N. and Frizinsky S. and Hagin D. and Ov | 2020 | | | No | | | | | | | | | | |
| Ostojic S.M. | 2021 | No | | | | | | | | | | | | |
| Taylor B.E.S. and Khandelwal P. and Rallo M.S. | 2020 | | | No | | | | | | | | | | |
| Frontera J.A. and Sabadia S. and Lalchan R. and | 2021 | | | No | | | | | | | | | | |
| Anonymous. | 2020 | No | | | | | | | | | | | | |
| Suppadungsuk S. and Jayanama K. and Sungka | 2020 | | | | | | | | | | | | | |
| Tay M.R.J. and Ong P.L. and Puah S.H. and Tha | 2020 | | | No | | | | | | | | | | |
| Anugwom C.M. and Aby E.S. and Debes J.D. | 2021 | No | | | | | | | | | | | | |
| Khoo B. and Tan T. and Clarke S.A. and Mills E.( | 2021 | | | No | | | | | | | | | | |
| Abrashkin K.A. and Zhang J. and Poku A. | 2021 | | No | | | | | | | | | | | |
| Chamorro-de-Vega E. and Rodriguez-Gonzalez | 2021 | | | No | | | | | | | | | | |
| Mason H. and Roberts R. and Raj A. | 2021 | | | | | | | | | | | | | |
| Crawford R.L. and Bolin E.H. and Prodhan P. and | 2021 | | | No | No | | | | | | | | | |
| Zheng Q.-N. and Xu M.-Y. and Zheng Y.-L. and W | 2020 | | | No | No | | | | | | | | | |
| Burks S.M. and Rosas-Hernandez H. and Alenja | 2021 | | | | | | | | | | | | | |
| Heming M. and Li X. and Rauber S. and Mausbe | 2021 | | | No | | | | | | | | | | |
| Zou X. and Fang M. and Huang J. | 2020 | No | | | | | | | | | | | | |
| Kingstone T. and Taylor A.K. and O'Donnell C.A. | 2020 | | | | | | | | | | | | | |
| Beauchamp L.C. and Finkelstein D.I. and Bush A | 2020 | No | | | | | | | | | | | | |
| McMahon D.E. and Gallman A.E. and Hruza G.J | 2021 | No | | | | | | | | | | | | |
| Bernini S. and Stasolla F. and Panzarasa S. and | 2020 | | No | | | | | | | | | | | |
| Venkatesan P. | 2021 | No | | | | | | | | | | | | |
| Moss B. and Mahajan K. and Bermel R. and Hell | 2020 | | No | | | | | | | | | | | |
| Spielmanns M. and Pekacka-Egli A.-M. and Cecc | 2021 | | | | | | | | | | | | | |
| Baig A.M. AO - Baig, Abdul Mannan; ORCID: http | 2020 | No | | | | | | | | | | | | |
| Khodjieva D.T. and Kazakov B.Sh. and Kadirovn | 2020 | No | | | | | | | | | | | | |
| Pun B.T. and Badenes R. and Heras La Calle G. | 2021 | | | | No | | | | | | | | | |
| Kostro K.B. and Gershengoren L. | 2021 | No | | | | | | | | | | | | |
| Grigoletto I. and Cavalheri V. and Lima F.F. and | 2020 | No | | | | | | | | | | | | |
| Vechorko V.I. and Evsikov E.M. and Baikova O.A | 2020 | | | | | | | | No | | | | | |
| Drapkina O.M. and Karpov O.E. and Loukyanov | 2020 | | | | | | | | No | | | | | |
| Myall K.J. and Mukherjee B. and Castanheira A. | 2021 | | | No | | | | | | | | | | |
| Manzo C. and Serra-Mestres J. and Isetta M. and | 2021 | No | | | | | | | | | | | | |
| Ashton J. | 2020 | No | | | | | | | | | | | | |
| Oda J. and Muguruma T. and Matsuyama S. and | 2020 | | | No | | | | | | | | | | |
| Alonso-Lana S. and Marquie M. and Ruiz A. and | 2020 | No | | | | | | | | | | | | |
| Shih E. and DiMaio J.M. and Squiers J.J. and Ba | 2021 | | | No | | | | | | | | | | |
| Li Y. and Zhao J. and Ma Z. and McReynolds L.S | 2021 | | No | | | | | | | | | | | |
| Wostyn P. | 2021 | No | | | | | | | | | | | | |
| Hittesdorf E. and Panzer O. and Wang D. and St | 2021 | | | No | | | | | | | | | | |
| Anker M.S. and Landmesser U. and von Haehlin | 2020 | No | | | | | | | | | | | | |
| Dekosky S.T. and Kochanek P.M. and Valadka A | 2021 | No | | | | | | | | | | | | |
| Cheng Y.-Y. and Chou W. AO - Cheng, Yuan-Yan | 2021 | No | | | | | | | | | | | | |
| Zheng H.-Y. and Xu M. and Yang C.-X. and Tian | 2020 | | | No | | | | | | | | | | |
| Pan P. and Pan R. | 2021 | No | | | | | | | | | | | | |
| Sivan M. and Taylor S. | 2020 | | | | | | | | | | | | | |
| Feng Z. and Zhao H. and Kang W. and Liu Q. an | 2020 | | | No | | | | | | | | | | |
| Addis S.G. and Nega A.D. and Miretu D.G. AO - | 2021 | | No | | | | | | | | | | | |
| Karimi L. and Sales C. and Crewther S.G. and W | 2020 | No | | | | | | | | | | | | |
| Higgins V. and Sohaei D. and Diamandis E.P. an | 2020 | No | | | | | | | | | | | | |
| Bachani S. and Arora R. and Dabral A. and Marv | 2020 | | | No | | | | | | | | | | |
| Pimlott N. and Agarwal P. and McCarthy L.M. and | 2020 | | | | No | | | | | | | | | |
| Zhang L. and Jiang Y. and Wei M. and Cheng B. | 2020 | | | No | | | | | | | | | | |
| Ribeiro D.E. and Oliveira-Giacomelli A. and Glas | 2020 | No | | | | | | | | | | | | |
| Yelin D. and Margalit I. and Yahav D. and Runold | 2020 | No | | | | | | | | | | | | |
| Saniasiaya J. and Kulasegarah J. AO - Saniasiay | 2021 | No | | | | | | | | | | | | |
| Canavera K.E. and Elliott D.A. | 2020 | No | | | | | | | | | | | | |
| Jianrong L. and Lyu H. and Yuan X. and An Y. ar | 2020 | | | | | | | No | | | | | | |
| Ferravante A.F. and Plazzi F. and Valentini I. and | 2020 | | | | | | | No | | | | | | |
| Bernardi E. and Plazzi F. and Ferravante A. and | 2020 | | | | | | | No | | | | | | |
| Behrendt A. and Akbarian-Tefaghi L. and Amani | 2020 | | | | | | | No | | | | | | |
| de Havenon A. and Ney J.P. and Callaghan B. ar | 2021 | | | No | | | | | | | | | | |
| Nath A. AO - Nath, Avindra; ORCID: http://orcid.c | 2020 | No | | | | | | | | | | | | |
| Valenzano A. and Scarinci A. and Monda V. and | 2020 | No | | | | | | | | | | | | |
| Meng L. and Dong D. and Li L. and Niu M. and B | 2020 | | | No | | | | | | | | | | |
| Lyons D. and Frampton M. and Naqvi S. and Do | 2020 | No | | | | | | | | | | | | |
| Ju C.-R. and Guan W.-J. and He J.-X. | 2020 | | No | | | | | | | | | | | |
| Jaffri A. and Jaffri U.A. | 2020 | No | | | | | | | | | | | | |
| Patel J. and Pujary D. and Kaur G. and Deodhar | 2020 | | No | | | | | | | | | | | |
| Baller E.B. and Hogan C.S. and Fusunyan M.A. | 2020 | No | | | | | | | | | | | | |
| Kubisiak K. and Dalrymple L.S. and Ofsthun N.J. | 2020 | | | No | | | | | | | | | | |
| Bustos A.E. and Connery M. and Becerra rivera | 2020 | | | No | | | | | | | | | | |
| Ahmed I. and Tumi A. | 2020 | No | | | | | | | | | | | | |
| Lokugamage A.U. and Bowen M.-A. and Blair J. | 2020 | No | | | | | | | | | | | | |
| Liu J. and Wang T. and Cai Q. and Sun L. and H | 2020 | | | No | | | | | | | | | | |
| Anonymous. | 2020 | | | | | | | | | | | | No | |
| Baptista A.F. and Baltar A. and Okano A.H. and | 2020 | No | | | | | | | | | | | | |
| Virgens I.P.A. and Santana N.M. and Lima S.C.V | 2020 | No | | | | | | | | | | | | |
| Banerjee D. and Viswanath B. AO - Banerjee, De | 2020 | No | | | | | | | | | | | | |
| Morley J.E. | 2020 | No | | | | | | | | | | | | |
| Salehi S. and Reddy S. and Gholamrezanezhad | 2020 | No | | | | | | | | | | | | |
| Puntmann V.O. and Carerj M.L. and Wieters I. ar | 2020 | | | No | | | | | | | | | | |
| Lindner D. and Fitzek A. and Brauninger H. and | 2020 | | | No | | | | | | | | | | |
| Anwar M.M. and Badawi A.M. and Eltablawy N.A | 2020 | No | | | | | | | | | | | | |
| Frutos-Reoyo E.J. and Cantalapiedra-Puentes E | 2020 | | | | | | | | No | | | | | |
| Razavi S.Z.E. and Hosseini M. and Azadvari M. | 2020 | No | | | | | | | | | | | | |
| Shah B.N. and Schlosshan D. and McConkey H. | 2020 | No | | | | | | | | | | | | |
| Levy J.M. | 2020 | No | | | | | | | | | | | | |
| Xiong G.L. and Atkin A. and Moquin K. and Canc | 2020 | | | | | | | | | | | | No | |
| Kholin A.A. and Zavadenko N.N. and Nesterovsk | 2020 | | | | | | | | No | | | | | |
| Figueira Goncalves J.M. and Garcia-Talavera I. a | 2020 | | | | | | | | No | | | | | |
| White P. | 2020 | No | | | | | | | | | | | | |
| Savage M. and Dooley L. and Waldron T. and Br | 2020 | | | | | | | No | | | | | | |
| Afrin L.B. and Weinstock L.B. and Molderings G. | 2020 | No | | | | | | | | | | | | |
| Lavery A.M. and Preston L.E. and Ko J.Y. and Cl | 2020 | | | No | | | | | | | | | | |
| Lyadov K.V. and Koneva E.S. and Polushkin V.G | 2020 | | | No | | | | | | | | | | |
| Koparal M. and Kurt E. and Altuntas E.E. and Do | 2020 | | | | No | | | | | | | | | |
| D'Ardes D. and Carrarini C. and Russo M. and D | 2020 | | | | No | | | | | | | | | |
| Basci S. and Ata N. and Altuntas F. and Yigenogl | 2020 | | | No | | | | | | | | | | |
| Ros Dopico L. and Tung-Chen Y. and Pilares Ba | 2020 | | | | | | | | No | | | | | |
| Pero A. and Ng S. and Cai D. | 2020 | No | | | | | | | | | | | | |
| Klitzman R.L. AO - Klitzman, Robert L.; ORCID: | 2020 | No | | | | | | | | | | | | |
| Li L. and Yu P. and Yang M. and Xie W. and Hua | 2020 | | | | | | | | | | | | | |
| Pizzirusso M. and Carrion-Park C. and Clark U.S | 2020 | | No | | | | | | | | | | | |
| Sun G. and Zhang Y. and Liao Q. and Cheng Y. | 2020 | | | No | | | | | | | | | | |
| Meyer T. and Posthumus E. | 2020 | | | | | | | | No | | | | | |
| Hong K. and Cao W. and Liu Z. and Lin L. and Z | 2020 | | | No | | | | | | | | | | |
| Carosi G. and Morelli V. and Del Sindaco G. and | 2020 | | | No | | | | | | | | | | |
| Baig A.M. AO - Baig, Abdul Mannan; ORCID: http | 2020 | No | | | | | | | | | | | | |
| Bourguignon A. and Beaulieu C. and Belkaid W. | 2020 | | | No | | | | | | | | | | |
| Zha L. and Xu X. and Wang D. and Qiao G. and | 2020 | | | No | | | | | | | | | | |
| Mancuso C.A. and Duculan R. and Jannat-Khah | 2020 | | No | | | | | | | | | | | |
| Tveito K. | 2020 | No | | | | | | | | | | | | |
| Mardani M. | 2020 | No | | | | | | | | | | | | |
| McClafferty B. and Umer I. and Fye G. and Kepk | 2020 | No | | | | | | | | | | | | |
| Bryson W.J. AO - Bryson, W. Jeff; ORCID: http:// | 2020 | No | | | | | | | | | | | | |
| Shrestha C. and Ghimire C. and Acharya S. and | 2020 | | No | | | | | | | | | | | |
| Harris B.H.L. and Zuhair M. and Di Giovannanto | 2020 | | | No | | | | | | | | | | |
| Xia P. and Wen Y. and Duan Y. and Su H. and Ca | 2020 | | | No | | | | | | | | | | |
| Iaccarino M.A. and Tenforde A.S. and Zafonte R. | 2020 | No | | | | | | | | | | | | |
| Merz L.E. and Sinnenberg L. and Gerhard-Herm | 2020 | | | No | | | | | | | | | | |
| Shi P. and Ren G. and Yang J. and Li Z. and Der | 2020 | | | | No | | | | | | | | | |
| Anonymous. | 2020 | No | | | | | | | | | | | | |
| Parnes N. and Tousant C. and Perrine J. and De | 2020 | | No | | | | | | | | | | | |
| Delahoy M.J. and Whitaker M. and O'Halloran A. | 2020 | | | | No | | | | | | | | | |
| Chen F. and Wang X.-D. and Zhu K.-K. and Hu J | 2020 | | | | No | | | | | | | | | |
| van de Kraats G. and van Veen S. | 2020 | | | | | | | | No | | | | | |
| Cirillo B. and Brachini G. and Crocetti D. and Sa | 2020 | | | No | | | | | | | | | | |
| Anonymous. | 2020 | No | | | | | | | | | | | | |
| Bellmann-Weiler R. and Burkert F. and Schwaige | 2020 | | | No | | | | | | | | | | |
| Canham L.J.W. and Staniaszek L.E. and Mortim | 2020 | | | | No | | | | | | | | | |
| Yelin D. and Wirtheim E. and Vetter P. and Kalil A | 2020 | No | | | | | | | | | | | | |
| McNeary L. and Maltser S. and Verduzco-Gutier | 2020 | No | | | | | | | | | | | | |
| Williams F.M.K. and Muirhead N. and Pariante C | 2020 | | | | | | | | | | | | No | |
| Deana C. and Verriello L. and Pauletto G. and C | 2020 | No | | | | | | | | | | | | |
| Moss B.P. and Mahajan K.R. and Bermel R.A. ar | 2020 | | No | | | | | | | | | | | |
| Kemp H.I. and Corner E. and Colvin L.A. | 2020 | No | | | | | | | | | | | | |
| Heneka M.T. and Golenbock D. and Latz E. and | 2020 | No | | | | | | | | | | | | |
| Wise J. | 2020 | No | | | | | | | | | | | | |
| Curci C. and Pisano F. and Negrini F. and De Sir | 2020 | No | | | | | | | | | | | | |
| Bossu P. and Toppi E. and Sterbini V. and Spalle | 2020 | No | | | | | | | | | | | | |
| Cohen G. and Russo M.J. and Campos J.A. and | 2020 | | No | | | | | | | | | | | |
| Chen P. and Zhang Y. and Wen Y. and Guo J. an | 2020 | | | | No | | | | | | | | | |
| Maillart E. and Papeix C. and Lubetzki C. and Ro | 2020 | | | No | | | | | | | | | | |
| Wilson B.A. and Betteridge S. and Fish J. | 2020 | No | | | | | | | | | | | | |
| Inoue H. and Jinno M. and Ohta S. and Kishino | 2020 | | | No | | | | | | | | | | |
| Rosen T. and Ferrante L.E. and Liu S.W. and Be | 2020 | No | | | | | | | | | | | | |
| Asserraji M. and Maoujoud O. and Belarbi M. and | 2020 | No | | | | | | | | | | | | |
| D'Andrea S. and Berardicurti O. and Berardicurti | 2020 | | | | No | | | | | | | | | |
| Carmo A. and Pereira-Vaz J. and Mota V. and M | 2020 | | | No | | | | | | | | | | |
| Grewal P. and Pinna P. and Hall J.P. and Dafer R | 2020 | | | No | | | | | | | | | | |
| Sivan M. and Halpin S. and Hollingworth L. and | 2020 | | No | | | | | | | | | | | |
| Shah S. and Danda D. and Kavadichanda C. and | 2020 | No | | | | | | | | | | | | |
| Bektas A. and Schurman S.H. and Franceschi C | 2020 | No | | | | | | | | | | | | |
| Pallanti S. and Grassi E. and Makris N. and Gas | 2020 | No | | | | | | | | | | | | |
| Lin L. and Luo S. and Qin R. and Yang M. and W | 2020 | | | No | | | | | | | | | | |
| Falvey J.R. and Ferrante L.E. | 2020 | No | | | | | | | | | | | | |
| Prakash A. | 2020 | No | | | | | | | | | | | | |
| Turolla A. and Rossettini G. and Viceconti A. and | 2020 | No | | | | | | | | | | | | |
| Greenhalgh T. and Knight M. and A'Court C. and | 2020 | No | | | | | | | | | | | | |
| Najjar S. and Najjar A. and Chong D.J. and Pram | 2020 | No | | | | | | | | | | | | |
| Candemir I. and Ergun P. | 2020 | No | | | | | | | | | | | | |
| Martin C. and Andres P. and Bullon A. and Villeg | 2020 | | No | | | | | | | | | | | |
| Calderon-Larranaga A. and Dekhtyar S. and Vet | 2020 | No | | | | | | | | | | | | |
| Ambrosino P. and Papa A. and Maniscalco M. ar | 2020 | No | | | | | | | | | | | | |
| Orsini A. and Corsi M. and Santangelo A. and Ri | 2020 | No | | | | | | | | | | | | |
| You J. and Zhang L. and Ni-jia-Ti M.-Y.-Y. and | 2020 | | | No | | | | | | | | | | |
| Del Bello A. and Marion O. and Sallusto F. and D | 2020 | | | No | | | | | | | | | | |
| Vitacca M. and Migliori G.B. and Spanevello A. a | 2020 | | | No | | | | | | | | | | |
| Leo F. and Wormanns D. and Grohe C. | 2020 | | | | | | | | No | | | | | |
| Radmard S. and Epstein S.E. and Roeder H.J. a | 2020 | | | No | | | | | | | | | | |
| Tadolini M. and Codecasa L.R. and Garcia-Garci | 2020 | | | No | | | | | | | | | | |
| Hascup E.R. and Hascup K.N. AO - Hascup, Erin | 2020 | No | | | | | | | | | | | | |
| Fraser E. | 2020 | No | | | | | | | | | | | | |
| Benger M. and Williams O. and Siddiqui J. and S | 2020 | | | | | No | | | | | | | | |
| Gaspardone C. and Meloni C. and Preda A. and | 2020 | | | No | | | | | | | | | | |
| Gan R. and Rosoman N.P. and Henshaw D.J.E. | 2020 | No | | | | | | | | | | | | |
| Perrin R. and Riste L. and Hann M. and Walther | 2020 | | | | | No | | | | | | | | |
| van Tilburg T.G. and Steinmetz S. and Stolte E. a | 2020 | | No | | | | | | | | | | | |
| Mills J.P. and Kaye K.S. and Mody L. | 2020 | No | | | | | | | | | | | | |
| Carpiniello B. and Tusconi M. and di Sciascio G. | 2020 | No | | | | | | | | | | | | |
| Levy J. and Leotard A. and Lawrence C. and Pa | 2020 | | | | No | | | | | | | | | |
| Rivera-Lillo G. and Torres-Castro R. and Fregon | 2020 | No | | | | | | | | | | | | |
| Aguilar-Guerra T.L. and Reed G. | 2020 | No | | | | | | | | | | | | |
| Baldacchino A. and Radfar S.R. and De Jong C. | 2020 | | No | | | | | | | | | | | |
| Troyer E.A. and Kohn J.N. and Hong S. | 2020 | No | | | | | | | | | | | | |
| Lopez M. and Bell K. and Annaswamy T. and Jue | 2020 | No | | | | | | | | | | | | |
| Mahase E. | 2020 | No | | | | | | | | | | | | |
| Zarrabian S. and Hassani-Abharian P. | 2020 | No | | | | | | | | | | | | |
| Cao Q. and Lu Z. | 2020 | | | No | | No | | | | | | | | |
| Guler A.A. and Ozturk M.A. AO - Guler, Aslihan A | 2020 | | | No | | | | | | | | | | |
| Bhaskar S. and Bradley S. and Israeli-Korn S. ar | 2020 | No | | | | | | | | | | | | |
| Vitti-Ruela B.V. and Dokkedal-Silva V. and Rosa | 2020 | No | | | | | | | | | | | | |
| Ling Y. and Xu S.-B. and Lin Y.-X. and Tian D. ar | 2020 | | | No | | | | | | | | | | |
| Yu P. and Wei Q. and He C. | 2020 | | | | | No | | | | | | | | |
| Escalon M.X. and Herrera J. | 2020 | | | | | | | | | | | | | |
| Fichera G. and Stramare R. and De Conti G. and | 2020 | | | No | | | | | | | | | | |
| Stam H.J. and Stucki G. and Bickenbach J. | 2020 | No | | | | | | | | | | | | |
| Gustavson A.M. and Danilovich M.K. and Lesser | 2020 | No | | | | | | | | | | | | |
| Tezcan M.E. and Dogan Gokce G. and Ozer R.S | 2020 | | | No | | | | | | | | | | |
| Beghi E. and Feigin V. and Caso V. and Santaluc | 2020 | No | | | | | | | | | | | | |
| Briguglio M. and Bona A. and Porta M. and Dell' | 2020 | No | | | | | | | | | | | | |
| Nakamura K. and Nakano H. and Naraba H. and | 2020 | No | | | | | | | | | | | | |
| Aldubaisi S. and Almudarra S. | 2020 | | No | | | | | | | | | | | |
| Piano C. and Di Stasio E. and Primiano G. and J | 2020 | | No | No | | | | | | | | | | |
| Abboud H. and Abboud F.Z. and Kharbouch H. a | 2020 | No | | | | | | | | | | | | |
| Paniz-Mondolfi A. and Bryce C. and Grimes Z. ar | 2020 | | | | | No | | | | | | | | |
| Wang Z. and Sun R. and Wang X. and Gu S. and | 2020 | | | | No | | | | | | | | | |
| Gombar S. and Chang M. and Hogan C.A. and Z | 2020 | | | No | | | | | | | | | | |
| Grabowski D.C. and Joynt Maddox K.E. | 2020 | No | | | | | | | | | | | | |
| Taylor S. and Asmundson G.J.G. | 2020 | No | | | | | | | | | | | | |
| Rana W. and Mukhtar S. AO - Rana, Waleed; OF | 2020 | No | | | | | | | | | | | | |
| Wang R. and Pan M. and Zhang X. and Han M. a | 2020 | | | No | | | | | | | | | | |
| Li T. and Ye Y. and Chen Y. and Pan J. AO - Che | 2020 | | | | No | | | | | | | | | |
| Brugliera L. and Spina A. and Castellazzi P. and | 2020 | No | | | | | | | | | | | | |
| Biamonte, Flavia and Botta, Cirino and Mazzitelli | 2021 | | | No | | | | | | | | | | |
| Comella, Phillip H and Gonzalez-Kozlova, Edgar | 2021 | | | | No | | | | | | | | | |
| Chun, Hyung J and Coutavas, Elias and Pine, Al | 2021 | | | No | | | | | | | | | | |
| Wise, Jacqui | 2021 | No | | | | | | | | | | | | |
| Lou, Jerry J and Movassaghi, Mehrnaz and Gord | 2021 | No | | | | | | | | | | | | |
| Atherton, Helen and Briggs, Tracy and Chew-Gra | 2021 | No | | | | | | | | | | | | |
| Wang, Julia Y and Zhang, Wei and Roehrl, Mich | 2021 | | | No | | | | | | | | | | |
| Vink, Mark and Vink-Niese, Alexandra | 2020 | | No | | | | | | | | | | | |
| Pisano, Francesca and Giachero, Alberto and Ru | 2020 | | No | | | | | | | | | | | |
| Walsh-Messinger, Julie and Manis, Hannah and | 2020 | | | | | | | No | | | | | | |
| Quicke, Kendra and Gallichote, Emily and Sexto | 2020 | | | | | | | No | | | | | | |
| Rubel, Kolin and Sharma, Dhruv and Campiti, Vi | 2020 | | | | No | | | | | | | | | |
| Ding, Qiyue and Shults, Nataliia V and Harris, Br | 2020 | | No | | | | | | | | | | | |
| Remsik, Jan and Wilcox, Jessica A and Babady, | 2020 | | | | No | | | | | | | | | |
| Suzuki, Emily and Sakai, Tomoko and Hoshino, | 2020 | | | No | | | | | | | | | | |
| Gupta, Aakriti and Madhavan, Mahesh V and Po | 2020 | | | No | | | | | | | | | | |
| Japan ECMOnet for COVID-19 | 2020 | | | No | | | | | | | | | | |
| Wilson, Clare | 2020 | No | | | | | | | | | | | | |
| Torres-Castro, Rodrigo and Solis-Navarro, Lilian | 2021 | No | | | | | | | | | | | | |
| Stefano, George B and Ptacek, Radek and Ptac | 2021 | No | | | | | | | | | | | | |
| Yang, Stephen Su and Lipes, Jed and Dial, Sand | 2020 | | | No | | | | | | | | | | |
| An, Ya-Wen and Yuan, Bo and Wang, Jian-Chun | 2021 | | | No | | | | | | | | | | |
| Sivan, Manoj and Taylor, Sharon | 2020 | No | | | | | | | | | | | | |
| Vilches-Moraga, A and Price, A and Braude, P ar | 2020 | | | No | | | | | | | | | | |
| Khatoonabadi, Ahmad Reza and Joanette, Yves | 2020 | No | | | | | | | | | | | | |
| Frenkel-Yosef, Maya and Maytles, Ruth and Shri | 2020 | | No | | | | | | | | | | | |
| McKee, Michael and Moran, Christa and Zazove | 2020 | No | | | | | | | | | | | | |
| Krawczyk, Noa and Fingerhood, Michael I and A | 2020 | No | | | | | | | | | | | | |
| Thai, Pham Quang and Toan, Do Thi Thanh and | 2020 | | | No | | | | | | | | | | |
| Johnson, Shawn F and Tiako, Max Jordan Ngue | 2020 | No | | | | | | | | | | | | |
| Broderick, Joseph P and Elm, Jordan J and Jani | 2020 | No | | | | | | | | | | | | |
| Madanes, Sharon B and Levenson-Palmer, Rose | 2020 | No | | | | | | | | | | | | |
| Varatharaj, Aravinthan and Thomas, Naomi and | 2020 | No | | | | | | | | | | | | |
| Cooper, Joseph J and Ross, David A AI - ORCID | 2020 | No | | | | | | | | | | | | |
| Su, Si and Cui, Huan and Wang, Tao and Shen, | 2020 | No | | | | | | | | | | | | |
| Innocenti, Pierluigi and Puzzella, Antonella and M | 2020 | | No | | | | | | | | | | | |
| Dani M. and Dirksen A. and Taraborrelli P. and T | 2021 | | | | | No | | | | | | | | |
| van Gassel R.J.J. and Bels J.L.M. and Raafs A. | 2021 | | | No | | | | | | | | | | |
| Estraneo A. and Ciapetti M. and Gaudiosi C. and | 2021 | | | No | | | | | | | | | | |
| Sher L. | 2021 | No | | | | | | | | | | | | |
| Saggini R. and Paolucci T. and Bramanti A. and | 2020 | | | | | | | | | | | | No | |
| Halpin S. and O'Connor R. and Sivan M. AO - Ha | 2021 | No | | | | | | | | | | | | |
| Zampogna E. and Migliori G.B. and Centis R. an | 2021 | | | No | | | | | | | | | | |
| Garcia J.C.H. and Montellano E.I.A. and Gonzale | 2020 | | | | | | | | No | | | | | |
| Liu J.W.T.W. and de Luca R.D. and Mello Neto H | 2020 | No | | | | | | | | | | | | |
| Steere H.K. and Polich G. and Silver J.K. and Ha | 2021 | | | No | | | | | | | | | | |
| Zarrougui W. and Bejaoui N. and Sedrati F. and | 2020 | | | | | | | | | | | | No | |
| Miori S. and Lassola S. and Gatti F.L. and Sanna | 2020 | | | | | | | | | | | | No | |
| Del Rio C. and Collins L.F. and Malani P. | 2020 | No | | | | | | | | | | | | |
| Taquet M. and Luciano S. and Geddes J.R. and | 2021 | | | | No | | | | | | | | | |
| Curci C. and Negrini F. and Ferrillo M. and Berg | 2021 | | | No | | | | | | | | | | |
| Soriano J.B. and Waterer G. and Penalvo J.L. ar | 2020 | No | | | | | | | | | | | | |
| Raveendran A.V. AO - Raveendran A.V.; ORCID: | 2021 | No | | | | | | | | | | | | |
| Tsivgoulis G. and Fragkou P.C. and Lachanis S. | 2021 | | | | | No | | | | | | | | |
| Galvan-Tejada C.E. and Herrera-Garcia C.F. and | 2020 | | | | No | | | | | | | | | |
| Al Chikhanie Y. and Verges S. and Herengt F. an | 2020 | | | | | | | | | | | | No | |
| Anonymous. | 2020 | No | | | | | | | | | | | | |
| Davido B. and Seang S. and Tubiana R. and de | 2020 | No | | | | | | | | | | | | |
| Kaiser R. | 2020 | No | | | | | | | | | | | | |
| Ferraro F. and Calafiore D. and Dambruoso F. ar | 2020 | | | | | No | | | | | | | | |
| Hasichaolu and Zhang X. and Li X. and Li D. | 2020 | | | No | | | | | | | | | | |
| Cicco S. and Vacca A. and Cittadini A. and Marra | 2020 | No | | | | | | | | | | | | |
| Zuin M. and Rigatelli G. and Zuliani G. and Ronc | 2020 | No | | | | | | | | | | | | |
| Hopkins C. and Burges Watson D.L. and Kelly C | 2020 | No | | | | | | | | | | | | |
| Zhang S. and Liu L. and Yang B. and Li R. and L | 2020 | | | | | | No | | | | | | | |
| Curci C. and Pisano F. and Bonacci E. and Cam | 2020 | | | | | | | | | | | | No | |
| Hao F. and Tam W. and Hu X. and Tan W. and Ji | 2020 | | | | No | | | | | | | | | |
| Rubin R. | 2020 | No | | | | | | | | | | | | |
| Romao V.C. and Oliveira-Ramos F. and Cruz-Ma | 2020 | | | | No | | | | | | | | | |
| Wijeratne T. and Crewther S. | 2020 | No | | | | | | | | | | | | |
| Belli. et al | 2020 | | | | No | | | | | | | | | |
| Carfi A. and Bernabei R. and Landi F. | 2020 | | | | | | | | | | | | | |
| Lindholm E. and Tonnessen T.I. | 2020 | No | | | | | | | | | | | | |
| Shui T.-J. and Li C. and Liu H.-B. and Chen X. ar | 2020 | | | No | | | | | | | | | | |
| Alcantara Montero A. and Pacheco de Vasconce | 2020 | | | | | | | | No | | | | | |
| Negrini F. and Ferrario I. and Mazziotti D. and Be | 2020 | | | | | No | | | | | | | | |
| Anonymous. | 2020 | | | | | | | | | | | | No | |
| Thornton J. | 2020 | No | | | | | | | | | | | | |
| Altmann, Daniel M and Boyton, Rosemary J | 2021 | No | | | | | | | | | | | | |
| van Dijk, Fons | 2021 | | | | | | | | No | | | | | |
| Anonymous | 2020 | | | | | | | | | | | | No | |
| Facharztmagazine, Redaktion | 2020 | | | | | | | | | | | | No | |
| Menon, Vikas and Padhy, Susanta Kumar | 2020 | No | | | | | | | | | | | | |
| Meyer, Thorsten and Posthumus, Elain and fur d | 2020 | | | | | | | | No | | | | | |
| Becker, Frank and Laake, Jon Henrik and Hofso, | 2020 | | | | | | | | No | | | | | |
| Anonymous | 2020 | | | | | | | | | | | | No | |
| Anonymous | 2020 | | | | | | | | | | | | No | |
| Cothran, Thomas P and Kellman, Suzanne and | 2020 | No | | | | | | | | | | | | |
| Al-Jahdhami I. and Al-Naamani K. and Al-Mawali | 2021 | No | | | | | | | | | | | | |
| Komaroff A.L. and Bateman L. | 2020 | No | | | | | | | | | | | | |
| Garg P. and Arora U. and Kumar A. and Wig N. A | 2021 | No | | | | | | | | | | | | |
| Rashid A. | 2020 | No | | | | | | | | | | | | |
| Boucas A.P. and Rheinheimer J. and Lagopoulos | 2020 | No | | | | | | | | | | | | |
| Deana C. and Verriello L. and Pauletto G. and C | 2020 | No | | | | | | | | | | | | |
| Pallanti S. and Grassi E. and Makris N. and Gas | 2020 | No | | | | | | | | | | | | |
| Greenhalgh T. and Knight M. and A'Court C. and | 2020 | No | | | | | | | | | | | | |
| Najjar S. and Najjar A. and Chong D.J. and Pram | 2020 | No | | | | | | | | | | | | |
| Orsini A. and Corsi M. and Santangelo A. and Ri | 2020 | No | | | | | | | | | | | | |
| Rolfe, Robert J and Smith, Colin M and Wolfe, C | 2021 | No | | | | | | | | | | | | |
| Klok, Frederikus A and Boon, Gudula J A M and | 2020 | No | | | | | | | | | | | | |
| Cui Y. and Zhang L. and Gao Q. and Hou Y. and | 2020 | | | | | | | | | | | | No | |
| Anonymous. | 2020 | | | | | | | | | | | | No | |
| Becker F. and Laake J.H. and Hofso K. | 2020 | | | | | | | | | | | | No | |
| Gonzalez F.A.I. and Dip J.A. and London S. | 2021 | | | | | | | | | | | | No | |
| Topeli A. and Halacli B. and Ortac Ersoy E. and | 2020 | | | | | | | | | | | | No | |
| Anonymous. | 2020 | | | | | | | | | | | | No | |
